# Analysis of Eligibility Criteria Clusters Based on Large Language Models for Clinical Trial Design

**DOI:** 10.1101/2024.10.08.24315075

**Authors:** Alban Bornet, Philipp Khlebnikov, Florian Meer, Quentin Haas, Anthony Yazdani, Boya Zhang, Poorya Amini, Douglas Teodoro

## Abstract

**Objectives:** Clinical trials (CTs) are essential for improving patient care by evaluating new treatments’ safety and efficacy. A key component in CT protocols is the study population defined by the eligibility criteria. This study aims to evaluate the effectiveness of large language models (LLMs) in encoding eligibility criterion information to support CT protocol design.

**Materials and Methods:** We extracted eligibility criterion sections, phases, conditions, and interventions from CT protocols available in the ClinicalTrials.gov registry. Eligibility sections were split into individual rules using a criterion tokenizer and embedded using LLMs. The obtained representations were clustered. The quality and relevance of the clusters for protocol design was evaluated through 3 experiments: intrinsic alignment with protocol information and human expert cluster coherence assessment, extrinsic evaluation through CT-level classification tasks, and eligibility section generation.

**Results:** Sentence embeddings fine-tuned using biomedical corpora produce clusters with the highest alignment to CT-level information. Human expert evaluation confirms that clusters are well-structured and coherent. Despite the high information compression, clusters retain significant CT information, up to 97% of the classification performance obtained with raw embeddings. Finally, eligibility sections automatically generated using clusters achieve 95% of the ROUGE scores obtained with a generative LLM.

**Conclusions:** We show that clusters derived from sentence-level LLM embeddings are effective in summarizing complex eligibility criterion data while retaining relevant CT protocol details. Clustering-based approaches provide a scalable enhancement in CT design that balances information compression with accuracy.

## INTRODUCTION

Clinical trials (CTs) are essential for advancing medical knowledge and improving patient care by systematically assessing the safety and efficacy of new treatments and interventions[1, 2]. A critical component of CTs is the protocol, which defines the eligibility criteria—specific characteristics that determine which participants are included or excluded from the study. Eligibility criteria balance the necessity to enroll enough participants with the need for a homogeneous study population and excluding individuals for whom the intervention could be unsafe[3].

Developing the eligibility section of the study protocol is a critical and complex task in CT design. On the one hand, under-restrictive criteria may lead to heterogeneous study populations, which can compromise the validity and efficacy of the CT results[4, 5] and increase the risk of adverse events[6]. On the other hand, overly restrictive criteria can significantly hinder participant recruitment[7]. It was shown that one CT in five fails to recruit enough participants, and most of them encounter delays because of the recruitment process[8, 9]. Historical analysis suggests that the number of participants is one of the main factors for clinical failure[10–12]. Moreover, restrictive criteria have a negative impact on the generalizability of CT results, which limits their range of application in future patients[13–15] and reduces their effectiveness in under-represented populations[16]. Hence, finding the optimal level of inclusivity and specificity in eligibility criteria is a major challenge in CT design.

Given this challenge, there is a growing research trend to develop data-driven solutions for optimizing and understanding CT data[17]. Publicly accessible databases of clinical studies, such as ClinicalTrial.gov[18], can be exploited by machine learning (ML) algorithms to analyze and predict CT data. ML research on eligibility criterion classification, recruitment prediction, patient-trial matching, or operational efficiency has met significant advancements in the last few decades (see[7, 19] for recent reviews).

### Background and Significance

Classic ML techniques have been applied to exploit the diverse features found in CT protocols. One branch explored the representation of free-text eligibility criteria into structured data using information extraction ML methods[20–22]. The goal is usually to help patient enrollment by automatically matching CTs based on patient medical records and estimate the number of eligible patients. Similarly, statistical modeling and various ML algorithms were used to predict recruitment rates and recruitment success in CTs[23–25]. Moreover, gradient boosting algorithms were trained to predict more specific features of operational efficiency in CTs, such as screen failure ratio, dropout ratio, or pre-enrollment delay[26]. Despite these successes, classic ML methods often struggle with the vast and varied nature of CTs and eligibility criteria, which requires continuous algorithm adaptation to their format.

In response to these limitations, recent advancements in natural language processing greatly improved the vectorial representation of unstructured text data (i.e., embeddings), such as the ones found in CT protocols, for classification and information extraction tasks. For example, pre-trained word embeddings were used to learn and predict eligibility for cancer CTs[27], and active learning was included in word embeddings to automatically classify eligibility criteria and enhance the efficiency of patient recruitment[28].

Attention-based models like BERT[29] and its large language model (LLM) variants better handle long-range dependencies and context-specific meanings, leading to improved performance with CT-protocol language. For example, BERT embeddings were used to match patients with appropriate CTs, and improved performance over classic language model baselines[30]. LLMs can be pre-trained with large databases from the general domain and further pre-trained with biomedical literature to generate embeddings that are more suited for clinical tasks, such as PubMedBERT[31] or BioBERT[32]. They can also be fine-tuned on more specific data domains or relevant tasks, which further improves the representation of CT data and patient records. For example, a study fine-tuned BioBERT on CT data and improved performance in a CT-retrieval task[33]. LLMs can also be flexibly integrated into complex architectures. For example, BERT was used to encode the content of leave nodes in a graph structure representing CT protocols for risk prediction and identified enrolment count as the main factor contributing to design-related risk[34, 35]. More recently, with the advent of generative LLMs, even more automated solutions were proposed to assist or improve CT design[36], patient-to-trial matching[37], and participant recruitment[38].

Recent work trained a BERT-like model with contrastive learning and rephrasing via generative LLMs to improve the quality of eligibility criterion embeddings[39]. Interestingly, clustering eligibility criteria was used to enhance the generation of negative sampling pairs. As semantically equivalent criteria can be described using different expressions, the current study focuses on evaluating the quality and relevance of the information available in semantic clusters. Relying on criterion clusters can identify common patterns and essential information across a large historical dataset of CTs while discarding irrelevant historical details. Moreover, clusters can be used to retrieve critical eligibility criteria which are more likely to be relevant and broadly applicable for a given set of phase(s), condition(s), and/or intervention(s).

To evaluate the amount and quality of the information available in criterion clusters, we performed three experiments.

- First, we intrinsically evaluated the alignment between generated clusters and information relevant to CT-protocol design by measuring mutual information scores between clusters and CT labels. Moreover, we directly assessed the coherence of generated eligibility criterion clusters using a human expert analysis.
- Second, we extrinsically evaluated information retained in clusters by using them as input features for ML models to classify CT-level outcomes and comparing the performance to using raw eligibility criterion embeddings.
- Third, we analyzed the effectiveness of semantic eligibility clusters to generate the eligibility criterion section of CT protocols and compared the results against those provided by a generative LLM prompted with CT information.

## MATERIALS AND METHODS

### Dataset

We extracted a dataset of eligibility criteria from the ClinicalTrials.gov registry[18], queried on 17.06.2024. Descriptive statistics for the dataset are available in Table A, Suppl. Inf. S6. From the database, we selected only interventional CTs:

- whose status was either completed or terminated,
- whose phase section included phase 1, 2, 3, or 4,
- and whose study started between 01.01.2000 and 01.06.2024.

Then, we split the criterion section of each CT into a list of individual criteria using a custom parsing algorithm based on sentence tokenization[40] and regular expression matching. Each criterion was associated with related CT-level information: phase(s), condition(s), intervention(s). Each condition and intervention were mapped to their corresponding MeSH[41] tree ID. Moreover, regular expression matching was used to identify, whenever possible, whether a criterion was exclusive or inclusive. The final raw dataset was written to a csv file in which each row contains an inclusion or exclusion criterion, the associated CT ID, matching condition and intervention MeSH IDs, and phase(s). The full dataset creation pipeline is shown in Figure 1A. The scripts to extract and build the dataset are available in our code repository.

**Figure 1.**
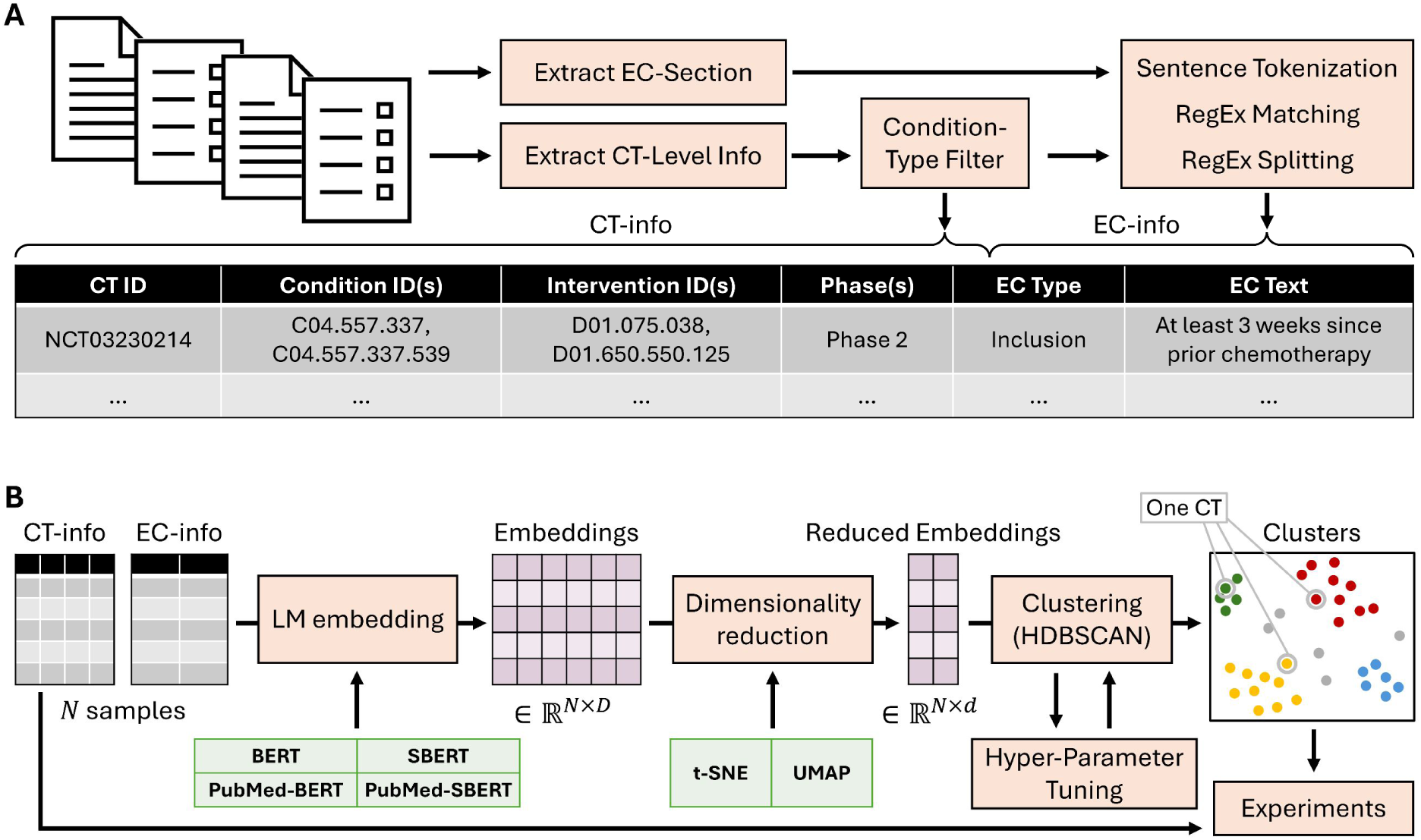
**A.** Eligibility criterion dataset generation. **B.** Cluster generation pipeline. Note: one CT is composed of several criteria and can be represented by a set of cluster IDs. **Acronyms.** EC: Eligibility Criterion; SBERT: Sentence-BERT.

### Cluster Generation

To cluster eligibility criteria based on LLM embeddings, we implemented a pipeline with three main steps – text embedding, dimensionality reduction, and clustering (Figure 1B). We adapted the BERTopic[42] package, which integrates most required functionalities. BERTopic combines BERT-like embeddings with a clustering algorithm and a class-based TF-IDF (which we did not use in our analysis), making it particularly suitable for organizing and summarizing the complex information found in CTs.

To embed eligibility criterion texts, we compared four different pre-trained language models: BERT[29], Sentence-BERT[43], a BERT model fine-tuned to produce sentence embeddings, PubMed-BERT[31], a BERT model further pre-trained on biomedical literature, and PubMed-Sentence-BERT[44], a PubMed-BERT model fine-tuned to produce sentence embeddings. Our goal was to determine the contribution of biomedical pre-training and the use of sentence embeddings in aligning criterion clusters with CT protocol information. For the token-level models, we used the output embedding of the [CLS] token as the numerical representation of a criterion. For the sentence-level models, we used the average of all non-[PAD] tokens.

Based on previous results[45, 46] showing that reducing dimensionality improves the performance of clustering algorithms with biomedical data, we implemented a dimensionality reduction step in our pipeline. We empirically selected t-SNE[47] with dimension 2 as this configuration has proven effective in creating qualitative clusters in our prior experiments[45, 46].

To improve the quality of the criteria clusters. we built a custom procedure on top of HDBSCAN[48] (BERTopic’s default) for the clustering step. First, clusters were generated from the reduced embeddings using HDBSCAN. Then, reduced embeddings of each “primary” cluster were fed to a new instance of HDBSCAN to attempt further “secondary” clustering. Primary clusters were kept intact in case the algorithm did not converge. The hyper-parameters of the clustering procedure were optimized using Optuna[49] (100 trials, TPE sampler[50]). The objective function was defined as follows:

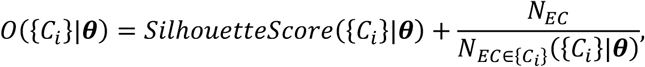

where {*C_i_*} is the result of the evaluated clustering procedure, ***θ*** the set of hyper-parameters, *N_EC_* the total number of eligibility criteria, and *N_EC_*_∈{*C*_*_i_*_}_ the number of criteria with an assigned cluster (HDBSCAN allows for unassigned samples). The set of hyper-parameters that were optimized by Optuna and their value ranges are described in Suppl. Inf. S1. To efficiently test many conditions and explore large hyper-parameter ranges, we used GPU-adapted versions of t-SNE, HDBSCAN, and Silhouette Score from the CuML suite[51].

We assessed cluster quality through three experiments summarized in Figure 2 and detailed in the next subsections. All experiments were run for different MeSH condition types: we filtered CT data to include only those with at least one condition matching C01 (Infections), C04 (Neoplasms), C14 (Cardiovascular Diseases), and C20 (Immune System Diseases).

**Figure 2.**
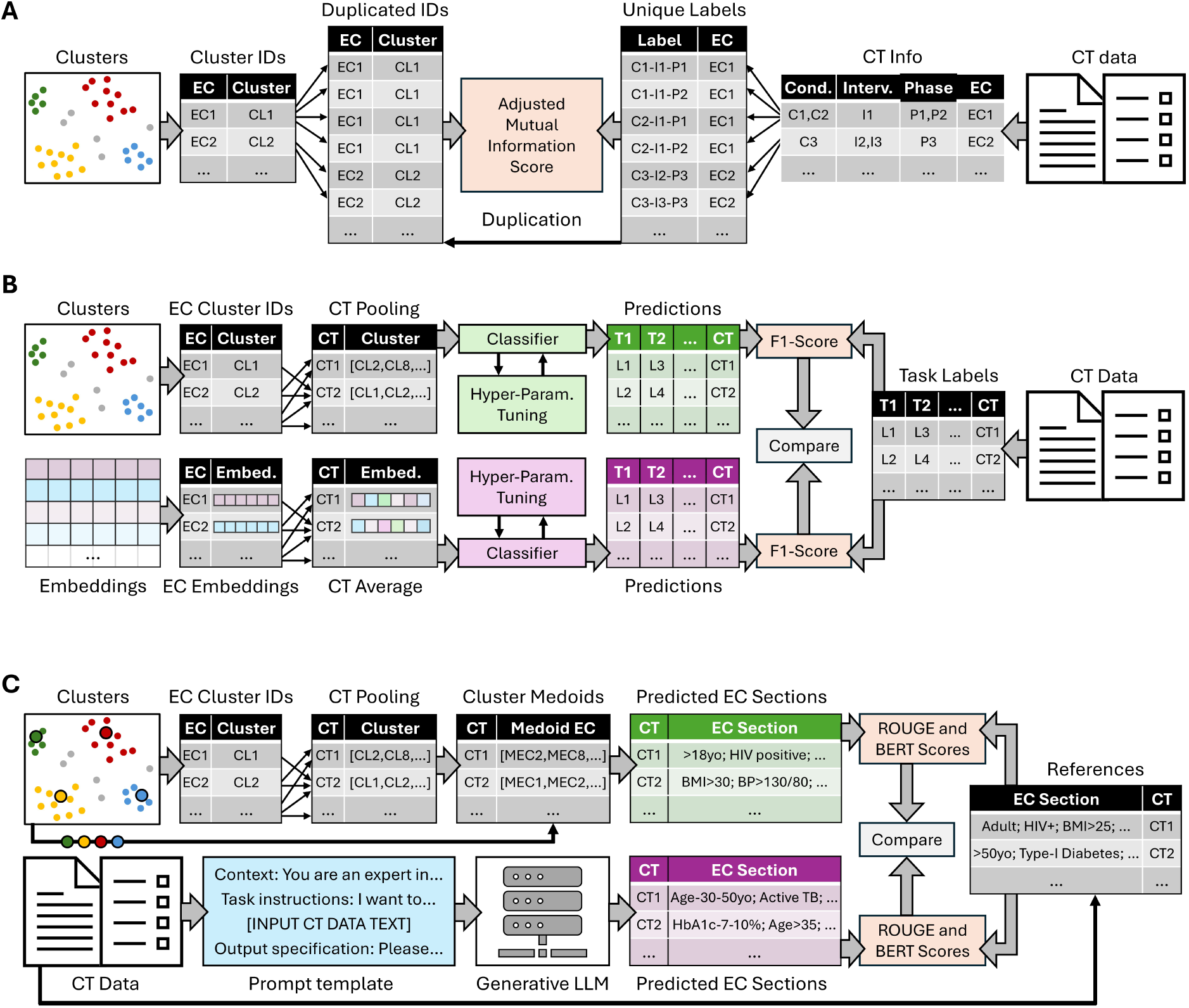
**A.** Experiment 1 – Alignment with CT-level information. **B.** Experiment 2 – CT-level classification. **C.** Experiment 3 – Eligibility section generation. **Acronyms.** EC – Eligibility Criterion; CL – Cluster; C – Condition; I – Intervention; P – Phase; T – Task; L – Label; MEC – Medoid EC.

### Experiment 1 – Intrinsic Evaluation – Alignment with CT-Level Information

To evaluate how eligibility clusters are aligned with the underlying structure of CTs, we measured the similarity of each cluster with CT-level information (Figure 2A). Each eligibility criterion was associated with a unique label, combining one CT phase, one condition, and one intervention. To reflect the required specificity of CTs, condition and intervention labels were derived from fourth-and third-level MeSH tree IDs, respectively. Eligibility criteria from CTs with several phases, conditions, or interventions, were duplicated for evaluation using each possible unique label combination. For example, if one criterion belonged to a CT with phases P_i_ and P_j_, intervention I_k_, and conditions C_m_ and C_n_, the corresponding duplicated samples would have the following labels: “P_i_-I_k_-C_m_”, “P_i_-I_k_-C_n_”, “P_j_-I_k_-C_m_”, and “P_j_-I_k_-C_n_”. We evaluated the alignment between clusters and CT-level information by computing the amount of mutual information between cluster IDs and the constructed labels. Given the high number of possible labels, we used the adjusted mutual information (AMI) score to correct the effect of random alignment between clusters and labels[52]. The resulting scores were compared across all models and condition types.

In addition, we asked a human expert to evaluate the quality of the clusters given a set of 1000 eligibility criteria. The clustering pipeline was used on CTs with conditions matching “C04” (Neoplasms). From the 50 largest clusters, 20 eligibility criteria closest to their cluster’s medoid were selected for evaluation. The expert evaluated whether the raw text of each eligibility criterion was correctly assigned to its respective cluster, considering the other criteria within the same cluster. Possible answers were “correct”, “not correct”, and “unclear”. The performance was computed as the number of correctly assigned samples over the total number of samples.

### Experiment 2 – Extrinsic Evaluation – CT-Level Classification

To extrinsically evaluate the quality of criterion clusters, we selected the best embedding model from Experiment 1 and used the generated clusters as input features for different CT-level classification tasks (Figure 2B). The goal of this experiment was to measure how much protocol information is retained by cluster IDs only, as compared to raw criterion embeddings. After splitting CTs into training (70%), validation (10%), and test (20%) sets, we trained regularized regression models (Lasso Regression[53], Ridge Regression[54], Elastic Net[55]) to predict CT-level outcomes such as phase, study duration, and enrolment count. For continuous outcomes, we created binary labels by splitting the data evenly, with 50% of instances in each class. Each CT was represented by a vector of length N, where N is the number of identified clusters. Each element in the vector indicated the frequency of a particular criterion cluster within that CT. Then, we compared performance to using the raw embeddings from which the clusters were derived as input features. Each CT was represented by the average of its eligibility criterion embeddings, i.e., by a vector of length D, where D is the dimension of the embedding model.

Classifier hyper-parameters were optimized with Optuna, setting validation F1-score as the objective function. The choice of the classification algorithm was part of the hyper-parameter set. Ranges for all hyper-parameters are described in Suppl. Inf. S2. For each condition, the algorithm with the best selected parameters was retrained using the training and validation splits and evaluated with F1-score computed on the test set.

### Experiment 3 – Eligibility Criterion Section Generation

To assess the applicability of semantic-equivalent eligibility clusters, we generated complete eligibility sections of study protocols only using clusters information (Figure 2C). We first filtered for specific condition types (C01, C04, C14, C20) and then randomly selected, for each condition type, 100 CTs that included at least one matching condition. The process used to select evaluated CTs was as follows. For each potentially selected CT, we retrieved all CTs from ClinicalTrials.gov that shared at least one common phase, intervention, and condition, excluding the evaluated CT itself. Filters for condition and intervention were initially applied using fourth- and third-level MeSH tree IDs. If fewer than 5,000 eligibility criteria were retrieved from the identified CTs, we relaxed the filters to third- and second-level MeSH IDs. If necessary, we further expanded to second- and first-level IDs. If insufficient criteria were still found, we moved to the next potentially selected CT. The final evaluation dataset included 400 CTs (i.e., 100 for each condition type), from which the eligibility section was extracted as the target reference, and where each CT was mapped to a set of at least 5,000 relevant eligibility criteria.

Using the best model from Experiment 1 and for each evaluated CT, we ran the clustering pipeline on the corresponding set of eligibility criteria to identify clusters expected to reflect relevant historical information. Then, we used the medoid and prevalence of the clusters to generate the eligibility section of the evaluated CT. For each cluster, we selected the criterion closest to the cluster medoid as a candidate. We then looped over selected criteria and added them to the generated section with a probability proportional to the corresponding cluster prevalence. We added criteria until the average number of eligibility criteria per CT, based on the condition type (21 for C01, 30 for C04, 21 for C14, and 24 for C20), was reached. We measured the quality of all generated sections with ROUGE[56] and BERTScore[57] metrics, using the eligibility section of the corresponding CTs in the evaluation set as references.

Finally, we compared the performance of the clustering method to a more expensive approach using a generative LLM, GPT-3.5-Turbo. The LLM was prompted with task instructions and output specifications, followed by the whole text of the evaluated CT, from which only the eligibility section was removed. The exact prompt template is shown in Suppl. Inf. S3. LLM-generated sections were evaluated with ROUGE and BERTscore metrics, using the same references as for the clustering method.

As a random baseline for both generation methods, we computed the ROUGE and BERTScore metrics against reference sections randomly sampled from the whole ClinicalTrials.gov dataset. This random baseline assesses each generation method’s lexicosemantic similarity with CT protocols in general. The difference between a method’s score and its random baseline measures how accurately the method captures the specific content of the target CT.

## RESULTS

In this section, we present the results of our three experiments, each aimed at evaluating different aspects of the information contained within eligibility criterion clusters. All statistical comparisons were done using paired t-tests, with a significance threshold set at an alpha level of 0.05.

### Experiment 1 – Intrinsic Evaluation – Alignment with CT-Level Information

Figure 3A shows the results of Experiment 1, in which clusters are evaluated intrinsically by computing the alignment between cluster IDs and labels relevant to CT-protocol design, using AMI. Clusters generated by the PubMed-Sentence-BERT model exhibit higher alignment with protocol information compared to other models, across all tested condition types. Moreover, both using sentence embeddings and using embeddings further pre-trained with biomedical data consistently improves cluster alignment. These results are in line with the visualization of eligibility clusters extracted from CTs matching condition type CO1 (Figure 3C – colors represent different clusters, and grey dots are samples with no assigned cluster). Clusters produced with sentence embedding are more separated and more samples find a cluster. Figure A-C, Suppl. Inf. S4, provide visualizations for other condition types.

**Figure 3.**
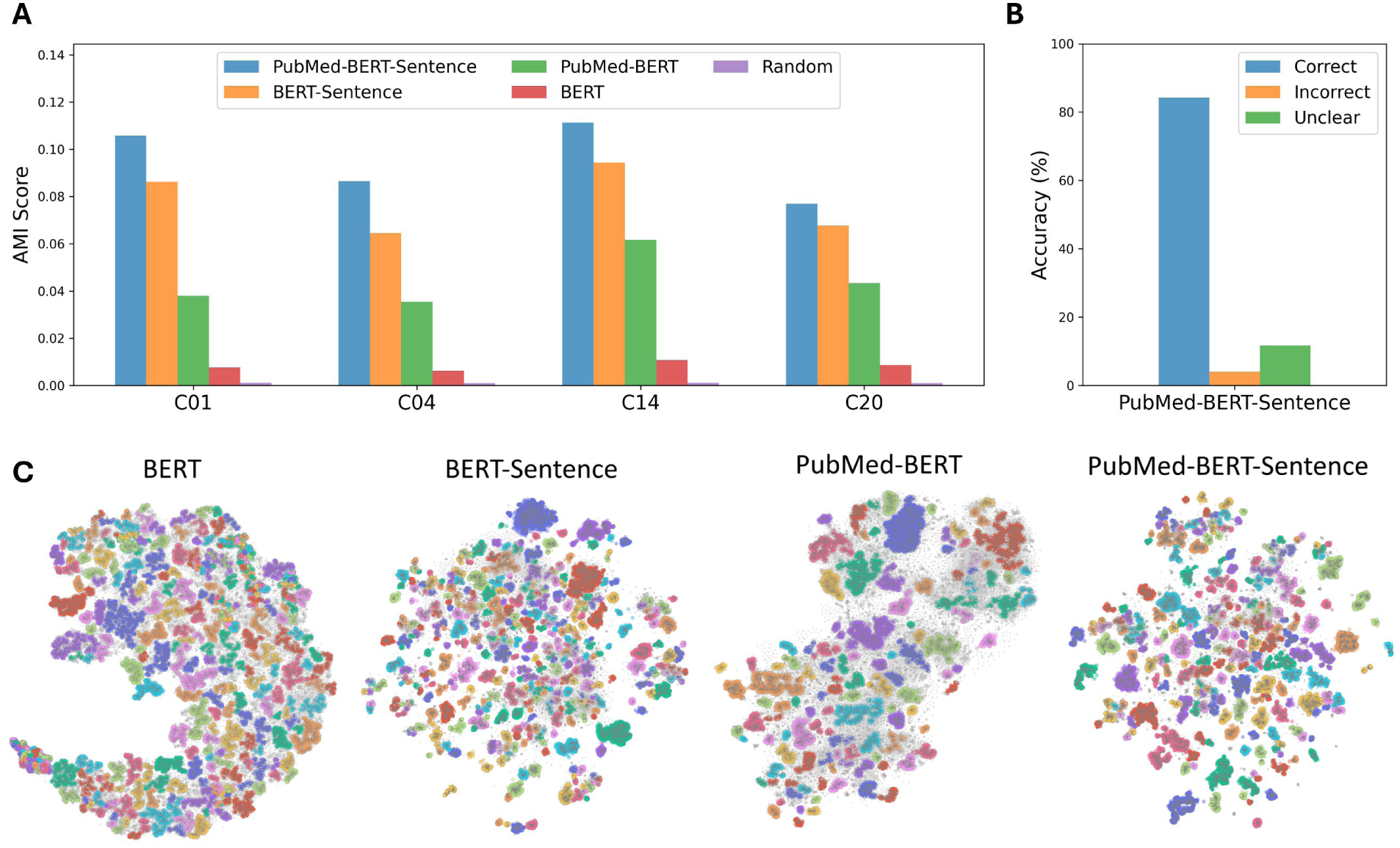
**A.** AMI scores for alignment with CT-protocol labels across different condition types. We visually added 0.001 to each score in order to see random performance, which was very close to 0. C01 – Infections; C04 – Neoplasms; C14 – Cardiovascular Diseases; C20 – Immune System Diseases. **B.** Human evaluation of cluster quality using PubMed-Sentence-BERT to embed eligibility criteria. **C.** Eligibility clusters extracted from CT protocols matching at least one condition starting with C01. Clusters are represented by different colors and grey dots are samples that were not assigned any cluster by the HDBSCAN algorithm. N_ECs_: 76,695; N_CTs_: 4,655; N_clusters_: {BERT: 245, BERT-Sentence: 200, PubMed-Bert: 310, PubMed-BERT-Sentence: 328}.

Figure 3B shows the results of the human expert cluster quality assessment for the best model identified in Figure 3A, i.e., PubMed-Sentence-BERT. Using 1000 criteria, the model obtains an accuracy score of 84.3%. This suggests that sentence-level embeddings are effective in producing coherent eligibility clusters.

### Experiment 2 – Extrinsic Evaluation – CT-Level Classification

Experiment 2 extrinsically evaluates the CT-level information encoded by the eligibility clusters. Using only the cluster IDs, different CT-level information is predicted. The results are compared to using raw embeddings as the upper bound, as they encode the maximum amount of eligibility information available to the clustering algorithm, and using random vectors as the lower bound, which do not encode any CT-level information. The random condition used uniform random vectors of the same dimensionality as for the cluster IDs condition. As for Experiment 1, this experiment was run separately for condition filters C01, C04, C14, and C20. For the phase classification task, all filtered samples were used. For the other classification tasks, the training, validation, and evaluation procedures were separated by phase. This means that there are 4 data points (each condition type filter) reported for the phase classification task, and 16 data points (4 condition type filters times 4 phase filters) reported for all other classification tasks.

Figure 4 shows macro-averaged F1-scores for different classification tasks and input features. For each classification task, we performed paired t-tests to compare the performance reached using cluster IDs to using raw embeddings, as well as using both input feature types to using random inputs (i.e., 12 comparisons in total). Performance using both cluster IDs and raw embeddings significantly outperforms the random baseline before and after Bonferroni correction (all task *p* < 0.004 for cluster IDs, and all task *p* < 0.002 for raw embeddings). Moreover, comparing performance using cluster IDs to using raw embeddings did not show any significant difference, before or after Bonferroni correction (*p*-values ranging from 0.135 for enrollment count classification to 0.626 for operational rate classification). Still, scores obtained with raw embedding are slightly higher than with cluster IDs. In terms of mean values, cluster IDs achieve on average scores that reach 97% of the scores obtained with raw embeddings, while the difference with the random baseline corresponds to 84% of the difference obtained between raw embeddings and the random baseline.

**Figure 4.**
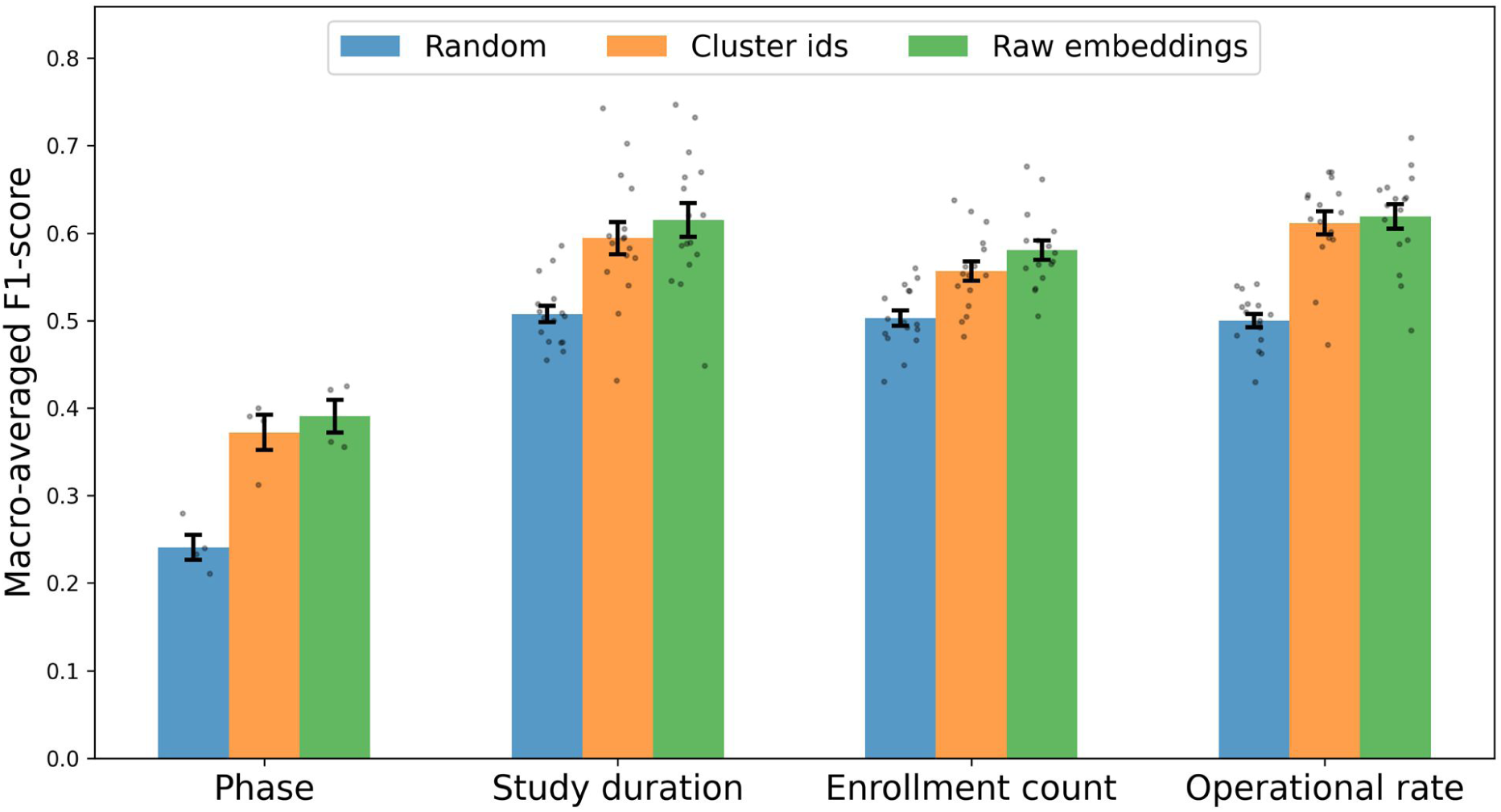
F1-scores for CT-level classification tasks comparing cluster-based features and raw embeddings. Error bars represent the standard error of the mean.

These results suggest that, despite significant compression compared to the raw embeddings, the clusters effectively retain the majority of essential information contained in CTs.

### Experiment 3 – Eligibility Criterion Section Generation

Experiment 3 evaluates the ability of clusters for the generation of eligibility criterion sections for new CT protocols. For each condition type (C01, C04, C14, C20), we randomly selected 100 CT protocols as the out-of-sample evaluation dataset, which we excluded from the analysis. We compared sections generated using eligibility clusters extracted from similar CT protocols to sections generated by prompting GPT-3.5-Turbo (more details in the methods section).

Figure 5 shows the average of the ROUGE-1, ROUGE-2, and ROUGE-L F1-scores, as well as BERTScore F_BERT_ (harmonic mean between recall *R*_BERT_ and precision *P*_BERT_, using SciBERT), computed over all samples of the evaluation dataset, for both methods and their associated random baselines. For both metrics, we performed paired t-tests to compare the cluster and generative LLM methods, as well as each method against its random baseline (i.e., 8 comparisons in total). Both the clustering and LLM methods significantly outperform their random baselines before and after Bonferroni correction, both in terms of average ROUGE F1-score (*p* < 0.001) and in terms of BERTScore F_BERT_ (*p* < 0.001). Moreover, the scores obtained with the clustering and LLM methods are significantly different from each other, both before and after Bonferroni correction (*p* < 0.001 for both metrics). This means that, when considering the 400 CTs from the evaluation dataset, the LLM method outperforms the clustering method. Still, in terms of mean values, the clustering method reaches 95% of the ROUGE F1-score and 98% of the BERTScore obtained with the generative LLM method (despite relying on an embedding model with O(10^3^) fewer parameters). Moreover, the difference between the average ROUGE F1-score obtained using the cluster method and its random baseline reaches 58% of the difference between the generative LLM method and its random baseline (and 65% for the BERTScore F_BERT_). Finally, the random baseline of the clustering method significantly outperforms the random baseline of the LLM method for the ROUGE F1-score, both before and after Bonferroni correction (*p* < 0.001).

**Figure 5.**
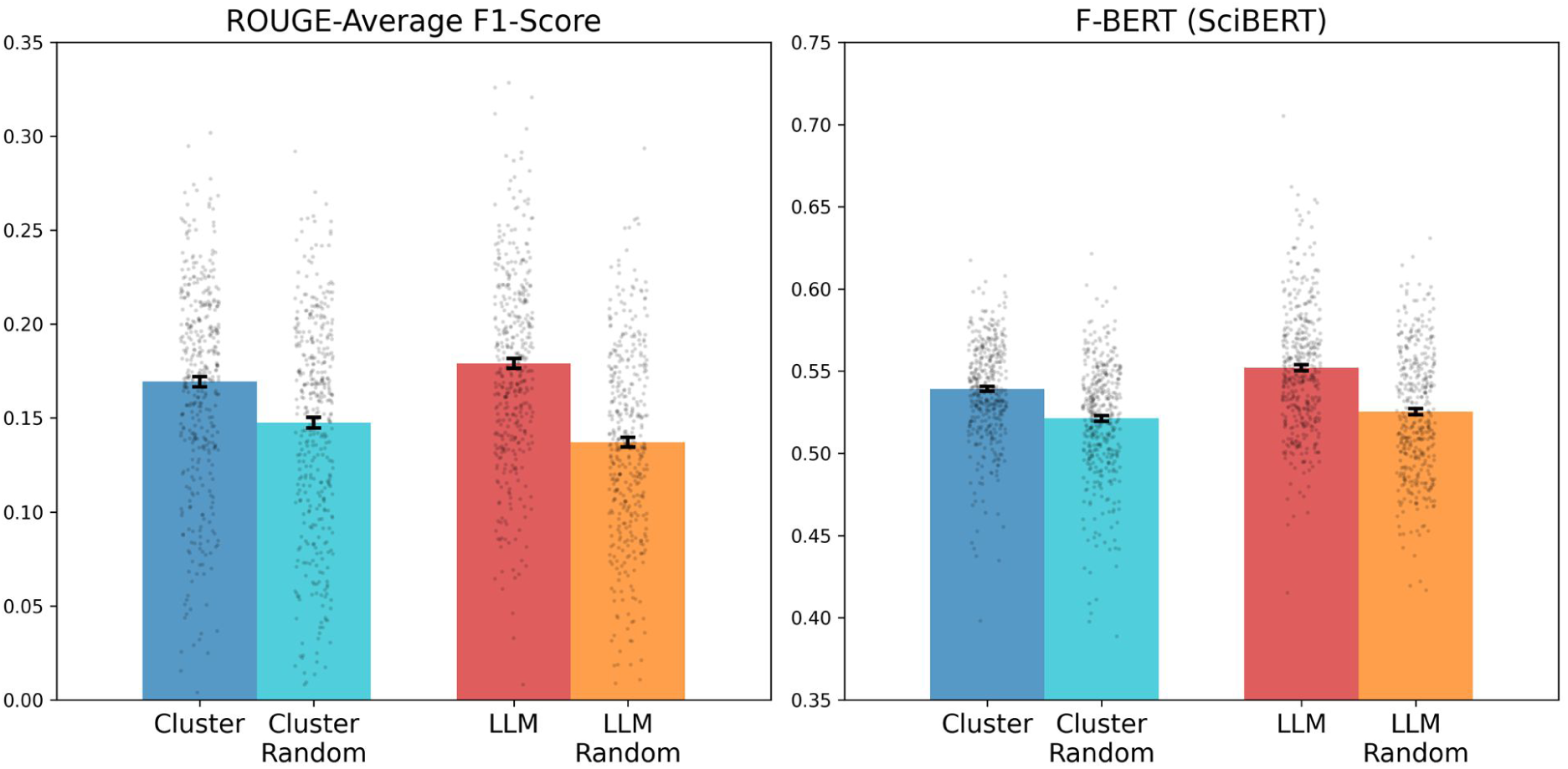
Average ROUGE F1-score and F_BERT_ (using SciBERT) for eligibility section generation using cluster medoids vs. generative LLM prompting, and corresponding random baselines. Error bars represent the standard error of the mean.

Suppl. Inf. S5 presents the same comparisons, stratifying by condition type (Figure D), and provides details for ROUGE-1, ROUGE-2, and ROUGE-L metrics as well as BERTScore, computed with different BERT models (Figure F for F1 scores, Figure G for recalls, Figure H for precisions). Finally, Figure E, Suppl. Inf. S5, shows a small, yet significant correlation between cluster quality, measured by silhouette score, and the metrics used in Figure 5 (*r* = 0.18 for average ROUGE F1-score, *r* = 0.23 for F_BERT_; *p* < 0.01 for both metrics). In contrast, no significant correlation is observed when using the random baseline scores (*p* = 0.23 for average ROUGE F1-score, *p* = 0.58 for F_BERT_).

## DISCUSSION

This study explores how eligibility criterion clusters extracted from CTs using encoder-based LLM embeddings align with CT protocol information and their effectiveness in summarizing complex CT data. The results obtained in our experiments show that clusters encode lexicosemantic eligibility criteria information offers insights that can be useful to improve the efficiency of CT design.

### Alignment of Clusters with Protocol-Design Information

The intrinsic evaluation of the clusters (Experiment 1) aims to directly evaluate their alignment with information relevant to protocol design. The AMI scores achieved by the different embedding models suggest that sentence-level embeddings fine-tuned on biomedical literature are the most effective in capturing relationships between different eligibility criteria and their corresponding CT characteristics. This highlights the importance of using tailored embeddings for extracting meaningful patterns from complex, unstructured clinical text data. Moreover, human expert evaluation confirms the coherence of the clusters generated with our best embedding model. The accuracy score (84.3%) achieved by the best model shows that clusters generated with sentence-level embeddings tailored for the biomedical domain align well with expert judgments.

### Retention of CT-Level Information in Clusters

The extrinsic evaluation of the clusters through CT-level classification tasks (Experiment 2) aims to assess how much relevant information is retained in compressed cluster representations. Although consistently lower, the cluster-based features achieve 97% of the performance obtained using raw embeddings. This suggests that semantic-equivalent eligibility criterion clusters, despite drastically compressing information (sparse, interpretable, 200- to 300-dimensional integer vectors instead of dense, non-interpretable, 768-dimensional floating-point vectors), retain most of the essential features present in raw embeddings. Cluster information offers a balance between data compression and the preservation of information which is crucial for scalable and efficient CT design. For example, when designing a new CT, screening all eligibility criteria from similar trials would be time-consuming and impractical, whereas cluster representations allow for rapid identification of relevant criteria while retrieving the most pertinent information. It should be noted that while raw embeddings are pooled using an average before being used as features for classification, their high dimensionality reduces the likelihood of significant information loss.

### Comparison with Large Language Model Outputs

When generating entire eligibility sections for CT protocols (Experiment 3), eligibility clusters derived from PubMed-Sentence-BERT embeddings perform reasonably well compared to outputs from a generative LLM (GPT-3.5-Turbo) prompted with nearly all information contained in the CT.

Using only the semantic eligibility cluster information achieves 95% of the ROUGE scores and 98% of the BERT scores reached by the generative LLM. While using only eligibility clusters to draft entire eligibility sections of CT protocols is likely not sufficient, the results of this experiment suggest that eligibility clusters contain information that is helpful for CT design. Our analysis was limited to cases where sufficient data could be extracted from ClinicalTrials.gov, given the set of phase(s), condition(s), and intervention(s) of the evaluated CT (i.e., at least 5,000 eligibility criteria). Expanding to larger or more diverse databases could mitigate this limitation.

It should be noted that the cluster random baseline, which assesses lexicosemantic similarity with CT protocols in general, significantly outperforms the LLM random baseline. This suggests that eligibility sections generated using cluster information are more aligned with the actual vocabulary of CTs, as our method directly pulls eligibility criteria from relevant trials based on cluster relevance. This is consistent with the findings shown in Figures G-H, Suppl. Inf. S5, where the cluster method generally outperforms the LLM method in terms of recall (sensitivity), while the LLM tends to show better precision.

Importantly, cluster quality, as measured by silhouette score, demonstrates a significant, yet small, correlation with the quality of the generated eligibility sections, as measured by both ROUGE and BERTScore (see Figure E, Suppl. Inf. S5). This suggests that when well-defined eligibility clusters are identified for a given clinical trial (CT), they are likely to provide valuable information for generating accurate eligibility sections. In contrast, when using random CT references to compute these metrics, no significant correlation is observed. This means that the improved performance of the cluster method over its random baseline is mostly due to the fact that cluster information helps generate sections that are more tailored to the specific requirements of the evaluated CT.

### Implications for CT Design

The findings of this study have significant implications for CT protocol design. Clustering eligibility criteria using encoder-based LLMs trained on biomedical corpora retains critical CT-level information while reducing the complexity of the data that clinical researchers need to screen. With a sufficiently large and diverse CT database and given that specific filtering (e.g., based on phase, conditions, and interventions) is provided, semantic-equivalent eligibility clusters could provide a compressed yet comprehensive overview of historically relevant eligibility criteria. This added information can help clinical researchers identify trends and patterns across various CTs, leading to better decision-making in the initial design phases. This approach ensures that no important information is overlooked during protocol design and that eligibility criteria are characterized by a wide range of pertinent historical data. Integrating such clustering methods into real-time data-driven tools could help the design process, ultimately improving the efficiency and success of CTs.

### Limitations

While our study presents interesting insights for CT protocol design, the metrics presented in our experiments remain relatively low. This is primarily due to the inherent challenge of comparing model outputs based on individual eligibility criteria to labels that are defined at the CT level. Moreover, further research is needed to explore the scalability of this approach across a broader range of CTs, conditions, and interventions. Future work could investigate the integration of clustering methods in data-driven tools, such as real-time updating of eligibility criteria based on new trial data becoming available. Finally, the potential and benefit of cluster information for CT design based on historical information remains to be demonstrated in a real use-case scenario.

## CONCLUSION

In conclusion, summarizing eligibility criterion information using cluster analysis based on LLMs provides a balance between information compression and CT-protocol relevance, particularly when using sentence-level embeddings from medically tailored language models. Our findings offer insights that could be used in advanced tools to enhance the design and efficiency of CT eligibility criteria.

## Supporting information

Suppl. Inf. S1

Suppl. Inf. S2

Suppl. Inf. S3

Suppl. Inf. S4

Suppl. Inf. S5

Suppl. Inf. S6

## Data Availability

The source data for this study is publicly available from ClinicalTrials.gov. The processed dataset generated and analyzed during the current study will be made available upon manuscript acceptance.

https://clinicaltrials.gov/

## AUTHOR CONTRIBUTIONS

A.B., D.T., and P.A. conceived and designed the study. A.B. was responsible for writing the manuscript, designing, implementing, and performing the experiments, as well as analyzing and visualizing the data. Q.H. contributed as the human expert for Experiment 1. P.K., F.M., A.Y., B.Z., P.A., and D.T. provided critical feedback on the manuscript. All authors reviewed and approved the final version of the manuscript.

## SUPPLEMENTARY MATERIAL

Supplementary material is available at Journal of the American Medical Informatics Association online.

