## Supplementary material for "Analysis of Eligibility Criteria Clusters Based on Large Language Models for Clinical Trial Design": Suppl. Inf. S1

### S1 – HDBSCAN Hyper-Parameters Fine-Tuned with Optuna

The hyper-parameters of HDBSCAN that were optimized with Optuna when clustering reduced eligibility criterion embeddings, and the range of possible values, were the following:

- For the first, “primary” cluster algorithm:
  - “max_cluster_size”: [0.01 * N, 0.1 * N]
  - “min_cluster_size”: [0.0 * N, 0.001 * N]
  - “min_samples”: min(128, [0.0 * N, 0.001 * N])
  - “alpha”: [0.1, 5.0]
  - “cluster_selection_method”: [“eom”, “leaf”]
- For all subsequent “secondary” clustering algorithms:
  - “max_cluster_size”: [0.1 * N, 1.0 * N]
  - “min_cluster_size”: [0.0 * N, 0.1 * N]
  - “min_samples”: min(128, [0.0 * N, 0.01 * N])
  - “alpha”: primary: [0.1, 5.0]
  - “cluster_selection_method”: [“eom”, ”leaf”]

Where N is the number of samples being clustered. Note that “min_samples” was capped to 128 to avoid out of memory errors.
