## Supplementary material for "Analysis of Eligibility Criteria Clusters Based on Large Language Models for Clinical Trial Design": Suppl. Inf. S2

### S2 – Classification Hyper-Parameters Fine-Tuned with Optuna

Model hyper-parameters for CT-level label classification algorithms were optimized with Optuna, with the following setup:

- algorithm taken from ["ridge", "lasso", "elasticnet"]
- if algorithm == "ridge":
    - scikit-learn algorithm = RidgeClassifier
    - alpha taken from (0.01, 10.0)
- if algorithm == "lasso":
    - scikit-learn algorithm = LogisticRegression
    - penalty = "l1"
    - solver taken from ["liblinear", "saga"]
    - C taken from (0.01, 10.0)
- if algorithm == "elasticnet":
    - scikit-learn algorithm = LogisticRegression
    - penalty = "elasticnet"
    - solver = "saga"
    - C taken from (0.01, 10.0)
    - l1_ratio taken from (0.0, 1.0)
