## Supplementary material for "Analysis of Eligibility Criteria Clusters Based on Large Language Models for Clinical Trial Design": Suppl. Inf. S3

### S3 – GPT-3.5 Turbo Prompt for Eligibility Section Generation

The prompt used to generate eligibility criterion sections by prompting GPT-3.5 Turbo with CT data was the following:

```
I have a clinical trial that includes the following information:

[CT_DATA_TEXT]

Based on the information above, generate the eligibility criteria section for this clinical trial.

Make sure the generated section includes [NUM_ELIGIBILITY_CRITERIA] eligibility criteria and has the following format:

Inclusion criteria:

<all inclusion criteria>

Exclusion criteria:

<all exclusion criteria>
```

Where [CT_DATA_TEXT] was replaced by the content of a CT from which only the eligibility criterion module was removed, and [NUM_ELIGIBILITY_CRITERIA] was replaced by the average number of eligibility criteria per CT in the run condition type (21 for C01, 30 for C04, 21 for C14, and 24 for C20).
