## Supplementary material for "Analysis of Eligibility Criteria Clusters Based on Large Language Models for Clinical Trial Design": Suppl. Inf. S4

### S4 – Cluster Comparisons for Other Condition Filters

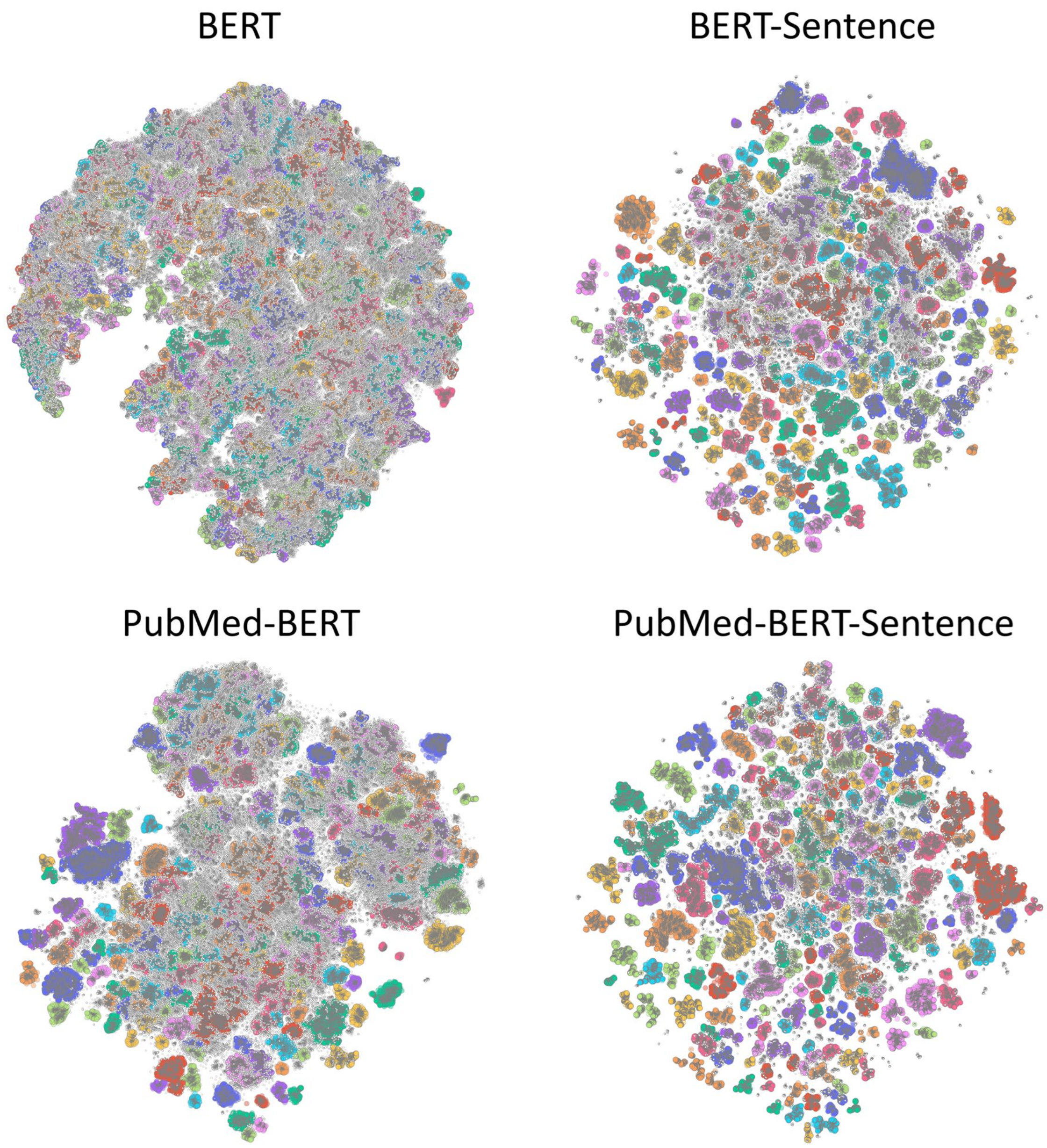


**Figure A.** Clusters extracted from CTs with at least one condition starting with C04 (Neoplasms). Colors represent different clusters, and grey dots are eligibility criteria with no defined cluster. $N_{ECs}$: 326,866; $N_{CTs}$: 14,985; $N_{clusters}$: {BERT: 358, BERT-Sentence: 247, PubMed-Bert: 242, PubMed-BERT-Sentence: 221}.

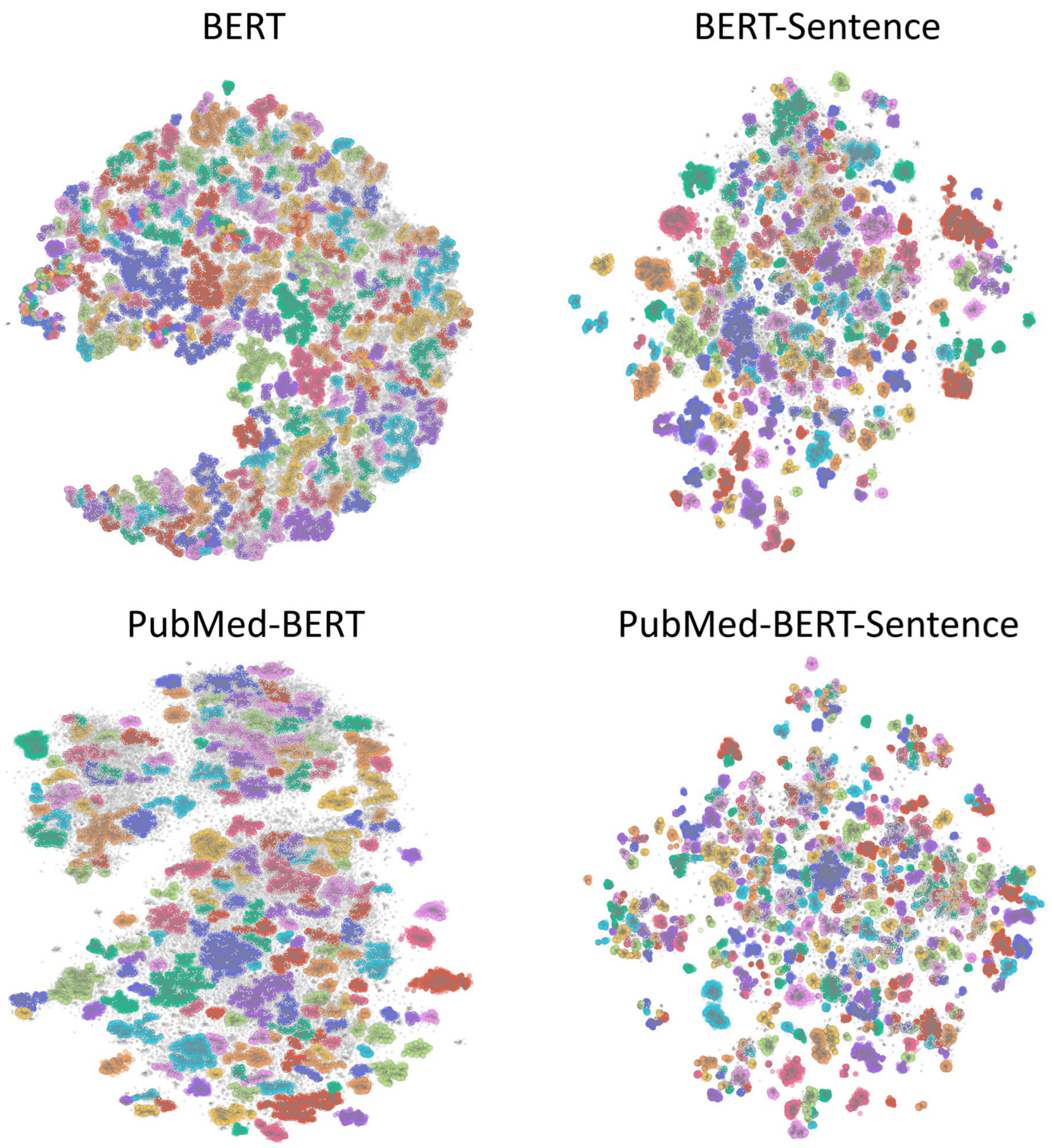


**Figure B.** Clusters extracted from CTs with at least one condition starting with C14 (Cardiovascular Diseases). Colors represent different clusters, and grey dots are eligibility criteria with no defined cluster. $N_{ECs}$: 87,948; $N_{CTs}$: 5,293; $N_{clusters}$: {BERT: 332, BERT-Sentence: 258, PubMed-Bert: 198, PubMed-BERT-Sentence: 454}.

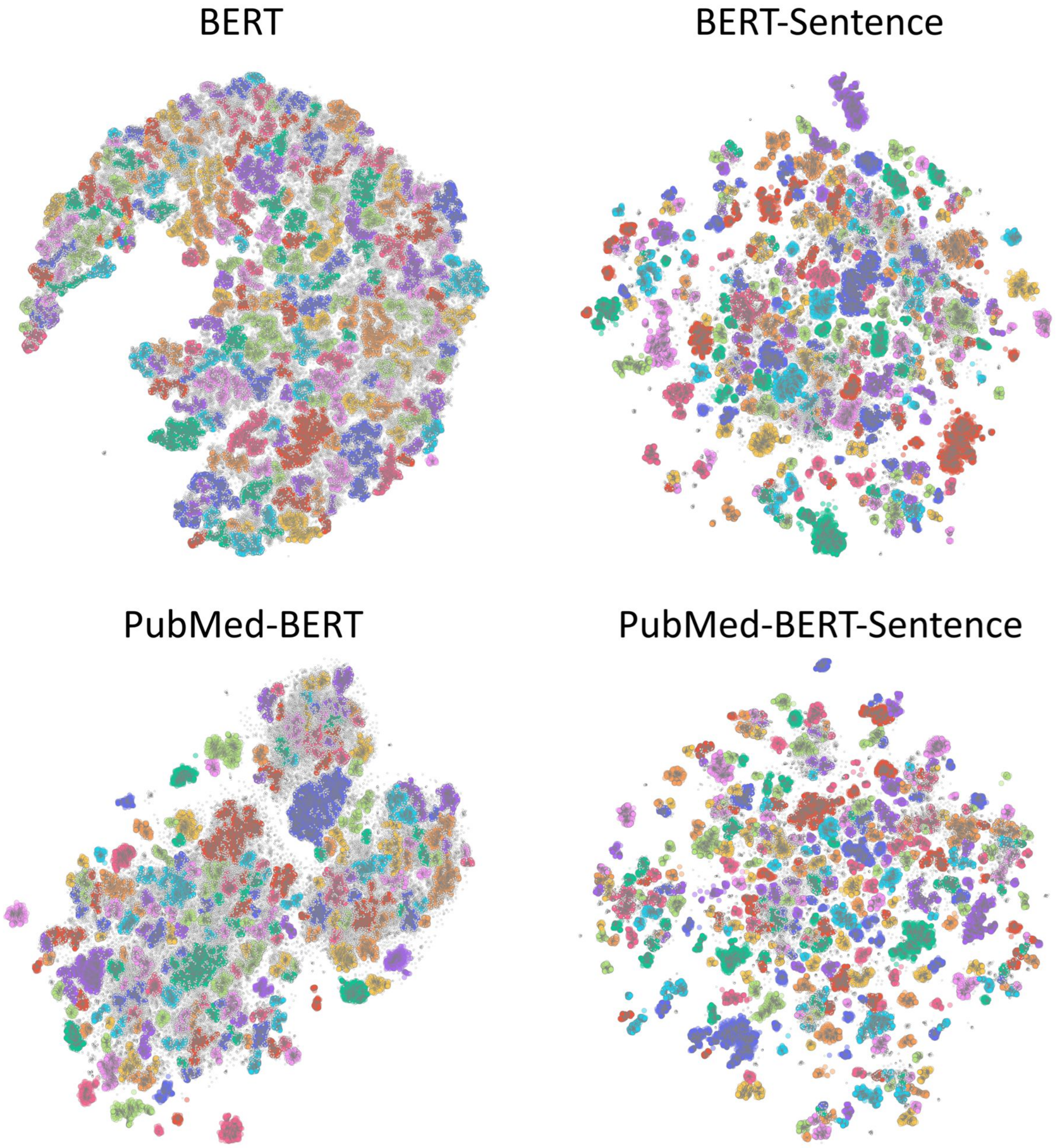


**Figure C.** Clusters extracted from CTs with at least one condition starting with C20 (Immune System Diseases). Colors represent different clusters, and grey dots are eligibility criteria with no defined cluster. $N_{ECs}$: 102,698; $N_{CTs}$: 4,873; $N_{clusters}$: {BERT: 252, BERT-Sentence: 302, PubMed-Bert: 232, PubMed-BERT-Sentence: 352}.
