## Supplementary material for "Analysis of Eligibility Criteria Clusters Based on Large Language Models for Clinical Trial Design": Suppl. Inf. S5

### S5 – Detailed Metrics for Eligibility Section Generation

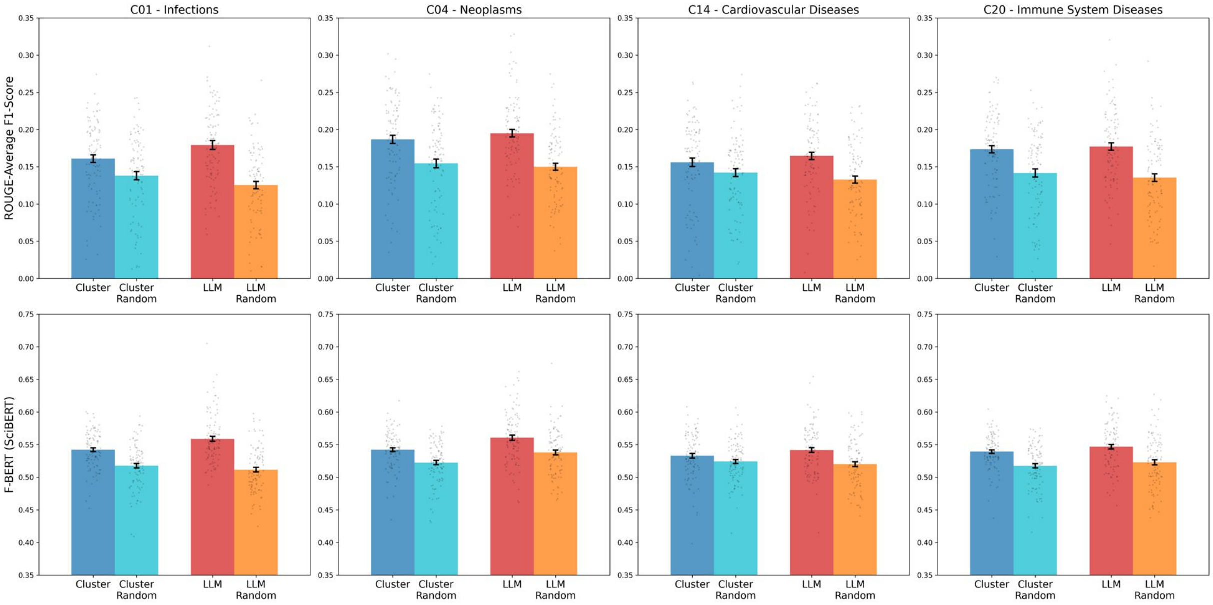


**Figure D.** Average ROUGE F1-score and $F_{BERT}$ (using SciBERT) for eligibility section generation using cluster medoids vs. generative LLM prompting, and corresponding random baselines, stratifying the evaluation dataset by CT condition type filter. Error bars represent the standard error of the mean.

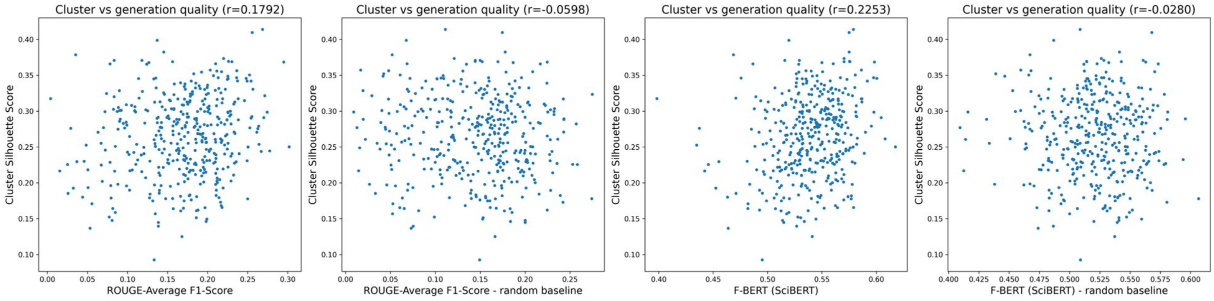


**Figure E.** Scatter plots of cluster quality, measured by silhouette score vs. eligibility section generation using cluster medoids, measured by Average ROUGE F1-score and BERTScore (using SciBERT). A significant correlation is observed for both metrics when comparing generated eligibility sections to the corresponding reference (first and third columns, $p < 0.001$ in both cases), whereas no significant correlation is observed when using random CT references to compute metrics ($p = 0.23$ for Average ROUGE F1-score, $p = 0.58$ for BERTScore).

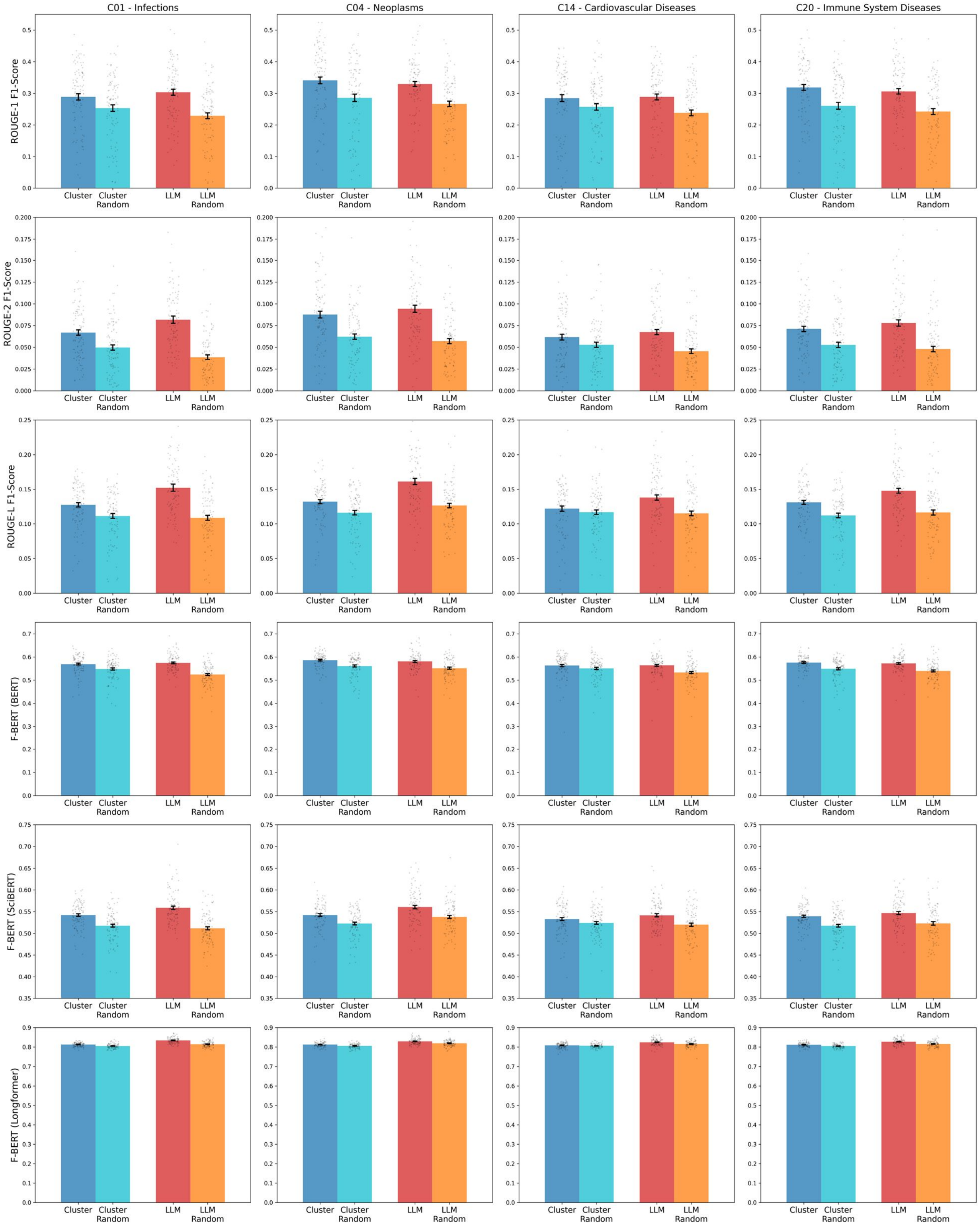


**Figure F.** ROUGE-1, ROUGE-2, and ROUGE-L, ROUGE-BERT, ROUGE-SciBERT, and Rouge-Longformer F1-scores comparing eligibility sections generated using cluster medoids vs LLM prompting, and corresponding shuffled baselines. Error bars represent the standard error of the mean.

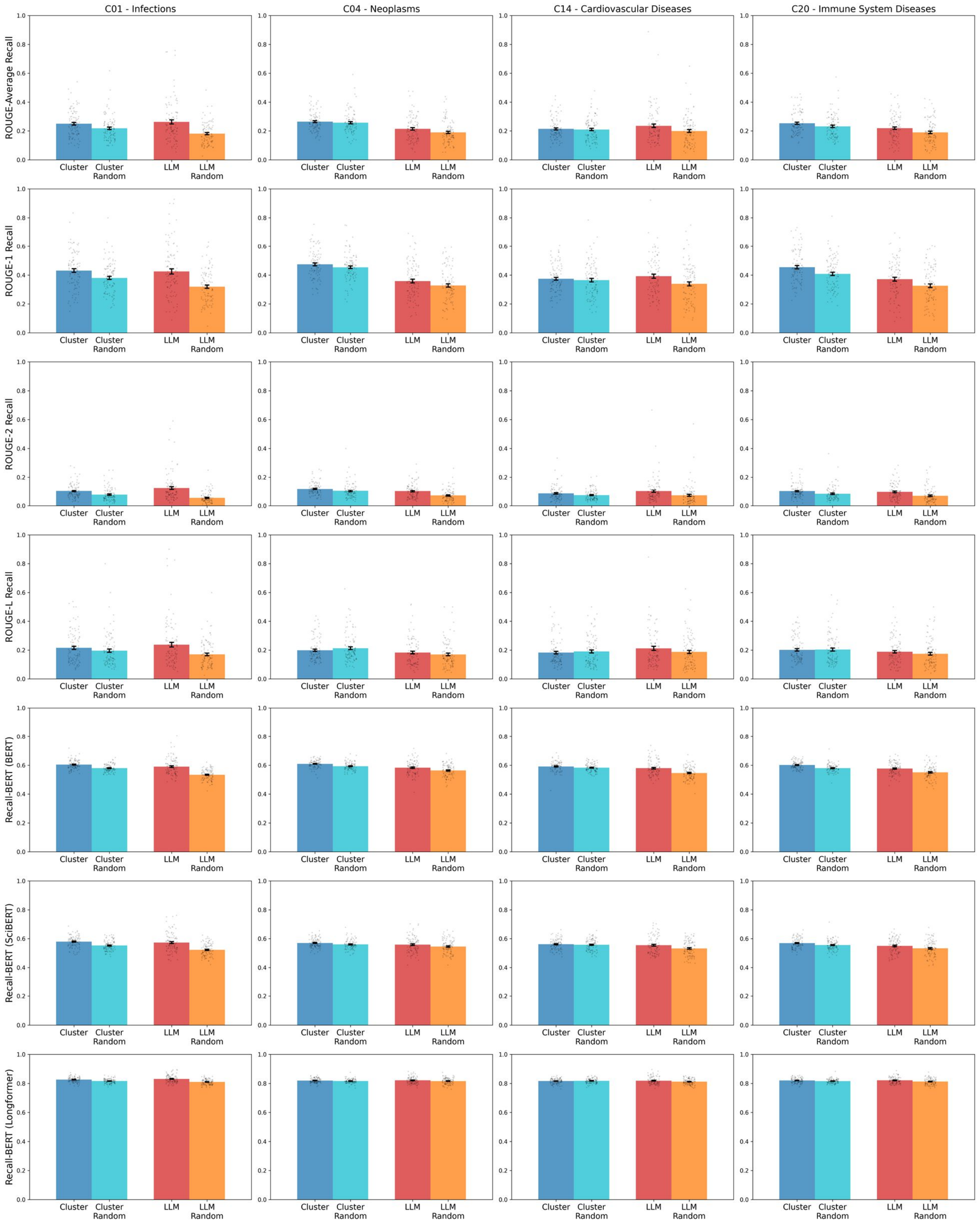


**Figure G.** ROUGE-1, ROUGE-2, and ROUGE-L, ROUGE-BERT, ROUGE-SciBERT, and Rouge-Longformer recalls comparing eligibility sections generated using cluster medoids vs LLM prompting, and corresponding random baselines, stratifying the evaluation dataset by CT condition type filter. Error bars represent the standard error of the mean.

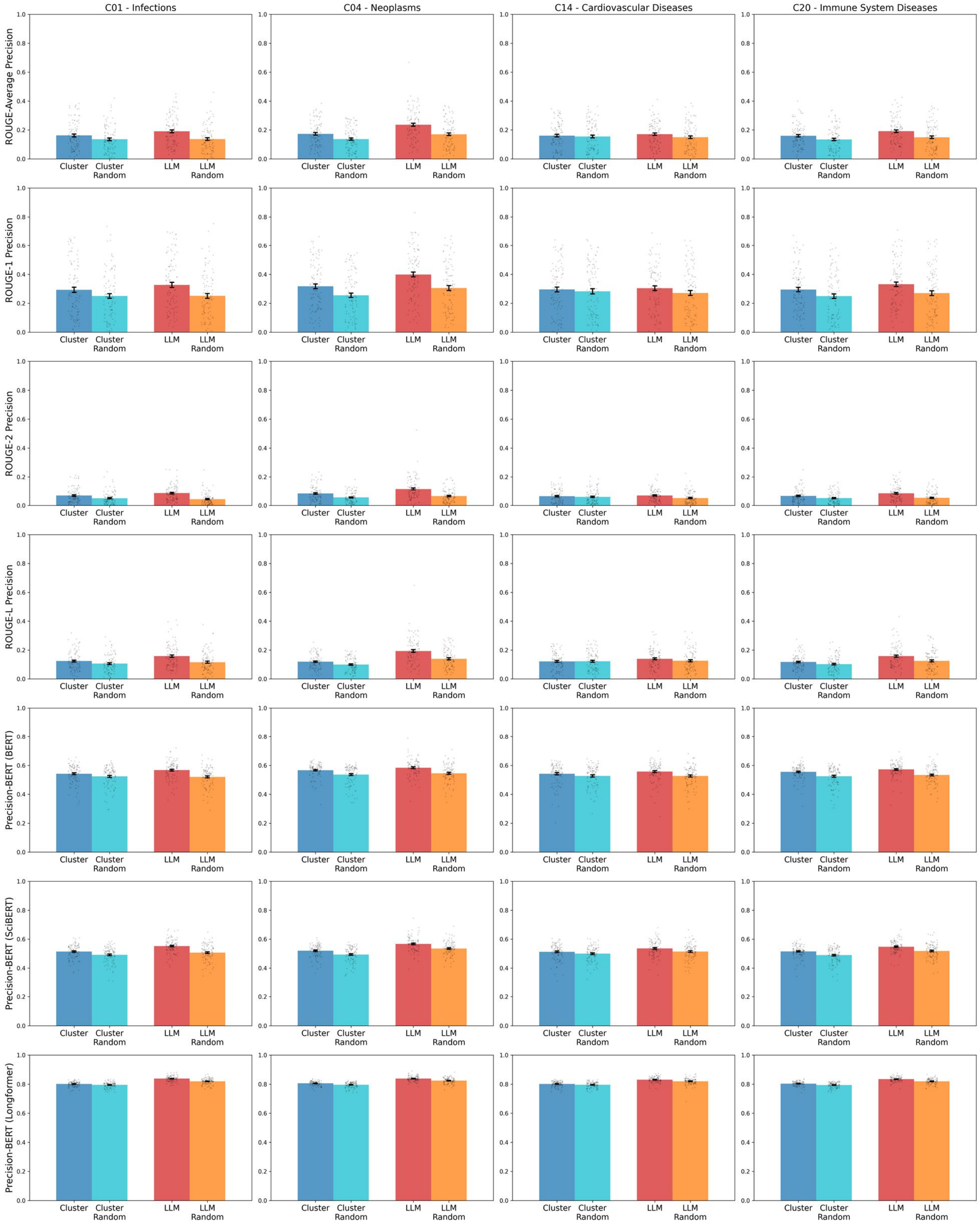


**Figure H.** ROUGE-1, ROUGE-2, and ROUGE-L, ROUGE-BERT, ROUGE-SciBERT, and Rouge-Longformer precisions comparing eligibility sections generated using cluster medoids vs LLM prompting, and corresponding random baselines, stratifying the evaluation dataset by CT condition type filter. Error bars represent the standard error of the mean.
