## Supplementary material for "Analysis of Eligibility Criteria Clusters Based on Large Language Models for Clinical Trial Design": Suppl. Inf. S6

### S6 – Dataset – Descriptive Statistics

| | | | | |
|---|---|---|---|---|
| ***Total*** | **Total EC Count** | **2071643** | **Total CT Count** | **98376** |
| | Phase 1 EC Count | 734574 (35.46%) | Phase 1 CT Count | 30223 (30.72%) |
| | Phase 2 EC Count | 878948 (42.43%) | Phase 2 CT Count | 36863 (37.47%) |
| | Phase 3 EC Count | 403262 (19.47%) | Phase 3 CT Count | 22653 (23.03%) |
| | Phase 4 EC Count | 245423 (11.85%) | Phase 4 CT Count | 16778 (17.05%) |
| | ECs per CT | 21.06 ± 15.59 | | |
| ***C01*** | **Total EC Count** | **195738 (9.45%)** | **Total CT Count** | **9434 (9.59%)** |
| | Phase 1 EC Count | 54995 (28.10%) | Phase 1 CT Count | 2044 (21.67%) |
| | Phase 2 EC Count | 75422 (38.53%) | Phase 2 CT Count | 3419 (36.24%) |
| | Phase 3 EC Count | 51759 (26.44%) | Phase 3 CT Count | 2799 (29.67%) |
| | Phase 4 EC Count | 31029 (15.85%) | Phase 4 CT Count | 1963 (20.81%) |
| | ECs per CT | 20.75 ± 15.60 | | |
| ***C04*** | **Total EC Count** | **740945 (35.77%)** | **Total CT Count** | **24998 (25.41%)** |
| | Phase 1 EC Count | 290313 (39.18%) | Phase 1 CT Count | 9136 (36.55%) |
| | Phase 2 EC Count | 456154 (61.56%) | Phase 2 CT Count | 14815 (59.26%) |
| | Phase 3 EC Count | 80967 (10.93%) | Phase 3 CT Count | 3387 (13.55%) |
| | Phase 4 EC Count | 12728 (1.72%) | Phase 4 CT Count | 816 (3.26%) |
| | ECs per CT | 29.64 ± 18.18 | | |
| ***C05*** | **Total EC Count** | **76898 (3.71%)** | **Total CT Count** | **4232 (4.30%)** |
| | Phase 1 EC Count | 14561 (18.94%) | Phase 1 CT Count | 677 (16.00%) |
| | Phase 2 EC Count | 29785 (38.73%) | Phase 2 CT Count | 1462 (34.55%) |
| | Phase 3 EC Count | 24983 (32.49%) | Phase 3 CT Count | 1464 (34.59%) |
| | Phase 4 EC Count | 13793 (17.94%) | Phase 4 CT Count | 942 (22.26%) |
| | ECs per CT | 18.17 ± 14.34 | | |
| ***C06*** | **Total EC Count** | **222708 (10.75%)** | **Total CT Count** | **9390 (9.55%)** |
| | Phase 1 EC Count | 70204 (31.52%) | Phase 1 CT Count | 2583 (27.51%) |
| | Phase 2 EC Count | 118326 (53.13%) | Phase 2 CT Count | 4546 (48.41%) |
| | Phase 3 EC Count | 39965 (17.95%) | Phase 3 CT Count | 2051 (21.84%) |
| | Phase 4 EC Count | 17715 (7.95%) | Phase 4 CT Count | 1130 (12.03%) |
| | ECs per CT | 23.72 ± 16.89 | | |
| ***C07*** | **Total EC Count** | **23422 (1.13%)** | **Total CT Count** | **1301 (1.32%)** |
| | Phase 1 EC Count | 4294 (18.33%) | Phase 1 CT Count | 189 (14.53%) |
| | Phase 2 EC Count | 13395 (57.19%) | Phase 2 CT Count | 663 (50.96%) |
| | Phase 3 EC Count | 5500 (23.48%) | Phase 3 CT Count | 378 (29.05%) |
| | Phase 4 EC Count | 2932 (12.52%) | Phase 4 CT Count | 249 (19.14%) |

| | | | | |
|---|---|---|---|---|
| | ECs per CT | 18.00 ± 14.24 | | |
| ***C08*** | **Total EC Count** | **221358 (10.69%)** | **Total CT Count** | **9454 (9.61%)** |
| | Phase 1 EC Count | 58335 (26.35%) | Phase 1 CT Count | 2069 (21.88%) |
| | Phase 2 EC Count | 108523 (49.03%) | Phase 2 CT Count | 4310 (45.59%) |
| | Phase 3 EC Count | 53320 (24.09%) | Phase 3 CT Count | 2601 (27.51%) |
| | Phase 4 EC Count | 22037 (9.96%) | Phase 4 CT Count | 1332 (14.09%) |
| | ECs per CT | 23.41 ± 16.75 | | |
| ***C09*** | **Total EC Count** | **26099 (1.26%)** | **Total CT Count** | **1249 (1.27%)** |
| | Phase 1 EC Count | 4651 (17.82%) | Phase 1 CT Count | 181 (14.49%) |
| | Phase 2 EC Count | 10835 (41.52%) | Phase 2 CT Count | 478 (38.27%) |
| | Phase 3 EC Count | 8337 (31.94%) | Phase 3 CT Count | 437 (34.99%) |
| | Phase 4 EC Count | 4480 (17.17%) | Phase 4 CT Count | 254 (20.34%) |
| | ECs per CT | 20.90 ± 15.82 | | |
| ***C10*** | **Total EC Count** | **192647 (9.30%)** | **Total CT Count** | **9706 (9.87%)** |
| | Phase 1 EC Count | 49811 (25.86%) | Phase 1 CT Count | 2077 (21.40%) |
| | Phase 2 EC Count | 86012 (44.65%) | Phase 2 CT Count | 4013 (41.35%) |
| | Phase 3 EC Count | 50585 (26.26%) | Phase 3 CT Count | 2843 (29.29%) |
| | Phase 4 EC Count | 27090 (14.06%) | Phase 4 CT Count | 1730 (17.82%) |
| | ECs per CT | 19.85 ± 14.51 | | |
| ***C11*** | **Total EC Count** | **48933 (2.36%)** | **Total CT Count** | **2914 (2.96%)** |
| | Phase 1 EC Count | 11450 (23.40%) | Phase 1 CT Count | 536 (18.39%) |
| | Phase 2 EC Count | 20773 (42.45%) | Phase 2 CT Count | 1176 (40.36%) |
| | Phase 3 EC Count | 13826 (28.25%) | Phase 3 CT Count | 882 (30.27%) |
| | Phase 4 EC Count | 9116 (18.63%) | Phase 4 CT Count | 657 (22.55%) |
| | ECs per CT | 16.79 ± 13.26 | | |
| ***C12*** | **Total EC Count** | **223205 (10.77%)** | **Total CT Count** | **9825 (9.99%)** |
| | Phase 1 EC Count | 71061 (31.84%) | Phase 1 CT Count | 2471 (25.15%) |
| | Phase 2 EC Count | 107746 (48.27%) | Phase 2 CT Count | 4203 (42.78%) |
| | Phase 3 EC Count | 41661 (18.66%) | Phase 3 CT Count | 2290 (23.31%) |
| | Phase 4 EC Count | 24899 (11.16%) | Phase 4 CT Count | 1757 (17.88%) |
| | ECs per CT | 22.72 ± 16.84 | | |
| ***C14*** | **Total EC Count** | **180401 (8.71%)** | **Total CT Count** | **9027 (9.18%)** |
| | Phase 1 EC Count | 42693 (23.67%) | Phase 1 CT Count | 1674 (18.54%) |
| | Phase 2 EC Count | 73691 (40.85%) | Phase 2 CT Count | 3266 (36.18%) |
| | Phase 3 EC Count | 45686 (25.32%) | Phase 3 CT Count | 2574 (28.51%) |
| | Phase 4 EC Count | 36050 (19.98%) | Phase 4 CT Count | 2280 (25.26%) |
| | ECs per CT | 19.98 ± 14.31 | | |
| ***C15*** | **Total EC Count** | **200329 (9.67%)** | **Total CT Count** | **7496 (7.62%)** |

| | | | | |
|---|---|---|---|---|
| | Phase 1 EC Count | 81186 (40.53%) | Phase 1 CT Count | 2626 (35.03%) |
| | Phase 2 EC Count | 116503 (58.16%) | Phase 2 CT Count | 4190 (55.90%) |
| | Phase 3 EC Count | 25141 (12.55%) | Phase 3 CT Count | 1241 (16.56%) |
| | Phase 4 EC Count | 6423 (3.21%) | Phase 4 CT Count | 441 (5.88%) |
| | ECs per CT | 26.72 ± 17.09 | | |
| ***C16*** | **Total EC Count** | **75839 (3.66%)** | **Total CT Count** | **4167 (4.24%)** |
| | Phase 1 EC Count | 21179 (27.93%) | Phase 1 CT Count | 1026 (24.62%) |
| | Phase 2 EC Count | 39208 (51.70%) | Phase 2 CT Count | 2007 (48.16%) |
| | Phase 3 EC Count | 18398 (24.26%) | Phase 3 CT Count | 1173 (28.15%) |
| | Phase 4 EC Count | 6575 (8.67%) | Phase 4 CT Count | 486 (11.66%) |
| | ECs per CT | 18.20 ± 13.28 | | |
| ***C17*** | **Total EC Count** | **194765 (9.40%)** | **Total CT Count** | **8471 (8.61%)** |
| | Phase 1 EC Count | 51157 (26.27%) | Phase 1 CT Count | 1870 (22.08%) |
| | Phase 2 EC Count | 101503 (52.12%) | Phase 2 CT Count | 4052 (47.83%) |
| | Phase 3 EC Count | 43765 (22.47%) | Phase 3 CT Count | 2243 (26.48%) |
| | Phase 4 EC Count | 16326 (8.38%) | Phase 4 CT Count | 999 (11.79%) |
| | ECs per CT | 22.99 ± 17.35 | | |
| ***C18*** | **Total EC Count** | **148559 (7.17%)** | **Total CT Count** | **8428 (8.57%)** |
| | Phase 1 EC Count | 39231 (26.41%) | Phase 1 CT Count | 2000 (23.73%) |
| | Phase 2 EC Count | 46602 (31.37%) | Phase 2 CT Count | 2400 (28.48%) |
| | Phase 3 EC Count | 43323 (29.16%) | Phase 3 CT Count | 2657 (31.53%) |
| | Phase 4 EC Count | 29753 (20.03%) | Phase 4 CT Count | 1911 (22.67%) |
| | ECs per CT | 17.63 ± 12.68 | | |
| ***C19*** | **Total EC Count** | **147259 (7.11%)** | **Total CT Count** | **7062 (7.18%)** |
| | Phase 1 EC Count | 42538 (28.89%) | Phase 1 CT Count | 1727 (24.45%) |
| | Phase 2 EC Count | 62238 (42.26%) | Phase 2 CT Count | 2535 (35.90%) |
| | Phase 3 EC Count | 34038 (23.11%) | Phase 3 CT Count | 1952 (27.64%) |
| | Phase 4 EC Count | 21654 (14.70%) | Phase 4 CT Count | 1373 (19.44%) |
| | ECs per CT | 20.85 ± 15.60 | | |
| ***C20*** | **Total EC Count** | **285730 (13.79%)** | **Total CT Count** | **11906 (12.10%)** |
| | Phase 1 EC Count | 99853 (34.95%) | Phase 1 CT Count | 3430 (28.81%) |
| | Phase 2 EC Count | 141240 (49.43%) | Phase 2 CT Count | 5493 (46.14%) |
| | Phase 3 EC Count | 53629 (18.77%) | Phase 3 CT Count | 2765 (23.22%) |
| | Phase 4 EC Count | 24627 (8.62%) | Phase 4 CT Count | 1451 (12.19%) |
| | ECs per CT | 24.00 ± 16.81 | | |
| ***C21*** | **Total EC Count** | **10 (0.00%)** | **Total CT Count** | **1 (0.00%)** |
| | Phase 1 EC Count | 0 (0.00%) | Phase 1 CT Count | 0 (0.00%) |
| | Phase 2 EC Count | 10 (100.00%) | Phase 2 CT Count | 1 (100.00%) |

| | | | | |
|---|---|---|---|---|
| | Phase 3 EC Count | 0 (0.00%) | Phase 3 CT Count | 0 (0.00%) |
| | Phase 4 EC Count | 0 (0.00%) | Phase 4 CT Count | 0 (0.00%) |
| | ECs per CT | 10.00 ± 0.00 | | |
| ***C22*** | **Total EC Count** | **1688 (0.08%)** | **Total CT Count** | **39 (0.04%)** |
| | Phase 1 EC Count | 973 (57.64%) | Phase 1 CT Count | 21 (53.85%) |
| | Phase 2 EC Count | 745 (44.14%) | Phase 2 CT Count | 18 (46.15%) |
| | Phase 3 EC Count | 42 (2.49%) | Phase 3 CT Count | 2 (5.13%) |
| | Phase 4 EC Count | 8 (0.47%) | Phase 4 CT Count | 1 (2.56%) |
| | ECs per CT | 43.28 ± 23.46 | | |
| ***C23*** | **Total EC Count** | **480769 (23.21%)** | **Total CT Count** | **24244 (24.64%)** |
| | Phase 1 EC Count | 113570 (23.62%) | Phase 1 CT Count | 4534 (18.70%) |
| | Phase 2 EC Count | 212171 (44.13%) | Phase 2 CT Count | 9563 (39.44%) |
| | Phase 3 EC Count | 118839 (24.72%) | Phase 3 CT Count | 6785 (27.99%) |
| | Phase 4 EC Count | 80829 (16.81%) | Phase 4 CT Count | 5496 (22.67%) |
| | ECs per CT | 19.83 ± 14.97 | | |
| ***C24*** | **Total EC Count** | **716 (0.03%)** | **Total CT Count** | **30 (0.03%)** |
| | Phase 1 EC Count | 182 (25.42%) | Phase 1 CT Count | 6 (20.00%) |
| | Phase 2 EC Count | 318 (44.41%) | Phase 2 CT Count | 12 (40.00%) |
| | Phase 3 EC Count | 243 (33.94%) | Phase 3 CT Count | 12 (40.00%) |
| | Phase 4 EC Count | 73 (10.20%) | Phase 4 CT Count | 3 (10.00%) |
| | ECs per CT | 23.87 ± 13.03 | | |
| ***C25*** | **Total EC Count** | **31114 (1.50%)** | **Total CT Count** | **1641 (1.67%)** |
| | Phase 1 EC Count | 8855 (28.46%) | Phase 1 CT Count | 408 (24.86%) |
| | Phase 2 EC Count | 14873 (47.80%) | Phase 2 CT Count | 772 (47.04%) |
| | Phase 3 EC Count | 5917 (19.02%) | Phase 3 CT Count | 338 (20.60%) |
| | Phase 4 EC Count | 4719 (15.17%) | Phase 4 CT Count | 305 (18.59%) |
| | ECs per CT | 18.96 ± 12.68 | | |
| ***C26*** | **Total EC Count** | **21110 (1.02%)** | **Total CT Count** | **1290 (1.31%)** |
| | Phase 1 EC Count | 4334 (20.53%) | Phase 1 CT Count | 206 (15.97%) |
| | Phase 2 EC Count | 9359 (44.33%) | Phase 2 CT Count | 508 (39.38%) |
| | Phase 3 EC Count | 5396 (25.56%) | Phase 3 CT Count | 352 (27.29%) |
| | Phase 4 EC Count | 4709 (22.31%) | Phase 4 CT Count | 387 (30.00%) |
| | ECs per CT | 16.36 ± 11.70 | | |
| ***D01*** | **Total EC Count** | **63773 (3.08%)** | **Total CT Count** | **2745 (2.79%)** |
| | Phase 1 EC Count | 20089 (31.50%) | Phase 1 CT Count | 697 (25.39%) |
| | Phase 2 EC Count | 32964 (51.69%) | Phase 2 CT Count | 1277 (46.52%) |
| | Phase 3 EC Count | 12303 (19.29%) | Phase 3 CT Count | 634 (23.10%) |
| | Phase 4 EC Count | 5660 (8.88%) | Phase 4 CT Count | 426 (15.52%) |

| | | | | |
|---|---|---|---|---|
| | ECs per CT | 23.23 ± 15.43 | | |
| ***D02*** | **Total EC Count** | **753350 (36.36%)** | **Total CT Count** | **35303 (35.89%)** |
| | Phase 1 EC Count | 229359 (30.45%) | Phase 1 CT Count | 8771 (24.84%) |
| | Phase 2 EC Count | 329738 (43.77%) | Phase 2 CT Count | 13019 (36.88%) |
| | Phase 3 EC Count | 155189 (20.60%) | Phase 3 CT Count | 8471 (24.00%) |
| | Phase 4 EC Count | 112984 (15.00%) | Phase 4 CT Count | 8020 (22.72%) |
| | ECs per CT | 21.34 ± 15.65 | | |
| ***D03*** | **Total EC Count** | **694545 (33.53%)** | **Total CT Count** | **32285 (32.82%)** |
| | Phase 1 EC Count | 222788 (32.08%) | Phase 1 CT Count | 8748 (27.10%) |
| | Phase 2 EC Count | 292863 (42.17%) | Phase 2 CT Count | 11459 (35.49%) |
| | Phase 3 EC Count | 142259 (20.48%) | Phase 3 CT Count | 7792 (24.14%) |
| | Phase 4 EC Count | 98694 (14.21%) | Phase 4 CT Count | 6804 (21.07%) |
| | ECs per CT | 21.51 ± 15.66 | | |
| ***D04*** | **Total EC Count** | **257170 (12.41%)** | **Total CT Count** | **12347 (12.55%)** |
| | Phase 1 EC Count | 72250 (28.09%) | Phase 1 CT Count | 2876 (23.29%) |
| | Phase 2 EC Count | 107162 (41.67%) | Phase 2 CT Count | 4465 (36.16%) |
| | Phase 3 EC Count | 61378 (23.87%) | Phase 3 CT Count | 3269 (26.48%) |
| | Phase 4 EC Count | 40605 (15.79%) | Phase 4 CT Count | 2781 (22.52%) |
| | ECs per CT | 20.83 ± 15.36 | | |
| ***D05*** | **Total EC Count** | **3972 (0.19%)** | **Total CT Count** | **248 (0.25%)** |
| | Phase 1 EC Count | 485 (12.21%) | Phase 1 CT Count | 25 (10.08%) |
| | Phase 2 EC Count | 1159 (29.18%) | Phase 2 CT Count | 67 (27.02%) |
| | Phase 3 EC Count | 1531 (38.54%) | Phase 3 CT Count | 89 (35.89%) |
| | Phase 4 EC Count | 1063 (26.76%) | Phase 4 CT Count | 85 (34.27%) |
| | ECs per CT | 16.02 ± 10.77 | | |
| ***D06*** | **Total EC Count** | **76750 (3.70%)** | **Total CT Count** | **4222 (4.29%)** |
| | Phase 1 EC Count | 18804 (24.50%) | Phase 1 CT Count | 981 (23.24%) |
| | Phase 2 EC Count | 26937 (35.10%) | Phase 2 CT Count | 1271 (30.10%) |
| | Phase 3 EC Count | 21345 (27.81%) | Phase 3 CT Count | 1219 (28.87%) |
| | Phase 4 EC Count | 15290 (19.92%) | Phase 4 CT Count | 1045 (24.75%) |
| | ECs per CT | 18.18 ± 13.62 | | |
| ***D08*** | **Total EC Count** | **33703 (1.63%)** | **Total CT Count** | **1476 (1.50%)** |
| | Phase 1 EC Count | 8078 (23.97%) | Phase 1 CT Count | 291 (19.72%) |
| | Phase 2 EC Count | 17556 (52.09%) | Phase 2 CT Count | 714 (48.37%) |
| | Phase 3 EC Count | 9105 (27.02%) | Phase 3 CT Count | 433 (29.34%) |
| | Phase 4 EC Count | 2749 (8.16%) | Phase 4 CT Count | 192 (13.01%) |
| | ECs per CT | 22.83 ± 15.46 | | |
| ***D09*** | **Total EC Count** | **86199 (4.16%)** | **Total CT Count** | **3741 (3.80%)** |

| | | | | |
|---|---|---|---|---|
| | Phase 1 EC Count | 25787 (29.92%) | Phase 1 CT Count | 975 (26.06%) |
| | Phase 2 EC Count | 41835 (48.53%) | Phase 2 CT Count | 1590 (42.50%) |
| | Phase 3 EC Count | 19895 (23.08%) | Phase 3 CT Count | 967 (25.85%) |
| | Phase 4 EC Count | 8635 (10.02%) | Phase 4 CT Count | 603 (16.12%) |
| | ECs per CT | 23.04 ± 16.30 | | |
| ***D10*** | **Total EC Count** | **53361 (2.58%)** | **Total CT Count** | **3132 (3.18%)** |
| | Phase 1 EC Count | 11977 (22.45%) | Phase 1 CT Count | 547 (17.46%) |
| | Phase 2 EC Count | 19733 (36.98%) | Phase 2 CT Count | 962 (30.72%) |
| | Phase 3 EC Count | 13026 (24.41%) | Phase 3 CT Count | 879 (28.07%) |
| | Phase 4 EC Count | 12560 (23.54%) | Phase 4 CT Count | 954 (30.46%) |
| | ECs per CT | 17.04 ± 13.05 | | |
| ***D12*** | **Total EC Count** | **342384 (16.53%)** | **Total CT Count** | **14800 (15.04%)** |
| | Phase 1 EC Count | 93143 (27.20%) | Phase 1 CT Count | 3474 (23.47%) |
| | Phase 2 EC Count | 179013 (52.28%) | Phase 2 CT Count | 6691 (45.21%) |
| | Phase 3 EC Count | 71592 (20.91%) | Phase 3 CT Count | 3691 (24.94%) |
| | Phase 4 EC Count | 36654 (10.71%) | Phase 4 CT Count | 2404 (16.24%) |
| | ECs per CT | 23.13 ± 16.86 | | |
| ***D13*** | **Total EC Count** | **92524 (4.47%)** | **Total CT Count** | **3627 (3.69%)** |
| | Phase 1 EC Count | 28312 (30.60%) | Phase 1 CT Count | 929 (25.61%) |
| | Phase 2 EC Count | 47792 (51.65%) | Phase 2 CT Count | 1740 (47.97%) |
| | Phase 3 EC Count | 17115 (18.50%) | Phase 3 CT Count | 804 (22.17%) |
| | Phase 4 EC Count | 10253 (11.08%) | Phase 4 CT Count | 541 (14.92%) |
| | ECs per CT | 25.51 ± 16.59 | | |
| ***D20*** | **Total EC Count** | **19868 (0.96%)** | **Total CT Count** | **780 (0.79%)** |
| | Phase 1 EC Count | 6024 (30.32%) | Phase 1 CT Count | 195 (25.00%) |
| | Phase 2 EC Count | 10614 (53.42%) | Phase 2 CT Count | 356 (45.64%) |
| | Phase 3 EC Count | 3172 (15.97%) | Phase 3 CT Count | 167 (21.41%) |
| | Phase 4 EC Count | 2626 (13.22%) | Phase 4 CT Count | 156 (20.00%) |
| | ECs per CT | 25.47 ± 17.77 | | |
| ***D23*** | **Total EC Count** | **76585 (3.70%)** | **Total CT Count** | **3703 (3.76%)** |
| | Phase 1 EC Count | 17794 (23.23%) | Phase 1 CT Count | 681 (18.39%) |
| | Phase 2 EC Count | 34561 (45.13%) | Phase 2 CT Count | 1497 (40.43%) |
| | Phase 3 EC Count | 19894 (25.98%) | Phase 3 CT Count | 1071 (28.92%) |
| | Phase 4 EC Count | 12090 (15.79%) | Phase 4 CT Count | 800 (21.60%) |
| | ECs per CT | 20.68 ± 15.65 | | |
| ***D25*** | **Total EC Count** | **3373 (0.16%)** | **Total CT Count** | **220 (0.22%)** |
| | Phase 1 EC Count | 339 (10.05%) | Phase 1 CT Count | 22 (10.00%) |
| | Phase 2 EC Count | 846 (25.08%) | Phase 2 CT Count | 55 (25.00%) |

| | | | | |
|---|---|---|---|---|
| | Phase 3 EC Count | 1307 (38.75%) | Phase 3 CT Count | 75 (34.09%) |
| | Phase 4 EC Count | 1010 (29.94%) | Phase 4 CT Count | 78 (35.45%) |
| | ECs per CT | 15.33 ± 10.01 | | |
| ***D26*** | **Total EC Count** | **33672 (1.63%)** | **Total CT Count** | **1886 (1.92%)** |
| | Phase 1 EC Count | 7845 (23.30%) | Phase 1 CT Count | 347 (18.40%) |
| | Phase 2 EC Count | 8070 (23.97%) | Phase 2 CT Count | 441 (23.38%) |
| | Phase 3 EC Count | 10831 (32.17%) | Phase 3 CT Count | 623 (33.03%) |
| | Phase 4 EC Count | 8847 (26.27%) | Phase 4 CT Count | 579 (30.70%) |
| | ECs per CT | 17.85 ± 13.52 | | |
| ***D27*** | **Total EC Count** | **79844 (3.85%)** | **Total CT Count** | **3711 (3.77%)** |
| | Phase 1 EC Count | 23521 (29.46%) | Phase 1 CT Count | 820 (22.10%) |
| | Phase 2 EC Count | 34641 (43.39%) | Phase 2 CT Count | 1334 (35.95%) |
| | Phase 3 EC Count | 17510 (21.93%) | Phase 3 CT Count | 975 (26.27%) |
| | Phase 4 EC Count | 12535 (15.70%) | Phase 4 CT Count | 917 (24.71%) |
| | ECs per CT | 21.52 ± 17.61 | | |

**Table A.** Descriptive statistics of the dataset extracted from ClinicalTrials.gov. EC – Eligibility Criterion. Statistics are stratified by MeSH condition ID types (C01 – C26) and MeSH intervention ID types (D01 – D27). Numbers add up to more than 100% because most CT protocols have several condition ID types, intervention ID types, or even several phases. Moreover, the total EC or CT counts reported in this table are smaller than the numbers reported in Figure 3, A, B, and C, since the ECs used to create plots in these figures went through an additional filtering step: selected ECs came from CTs with at least one MeSH condition ID and one MeSH intervention ID with enough depth to produce the labels for experiment 1 (i.e., 3 levels for condition, 4 levels for intervention).
